# *TTN* truncating variants in hiPSI exons show high penetrance for cardiomyopathy in carriers with atrial fibrillation

**DOI:** 10.1101/2022.06.06.22276058

**Authors:** Kelly M. Schiabor Barrett, Elizabeth T. Cirulli, Alexandre Bolze, Chris Rowan, Gai Elhanan, Joseph J. Grzymski, William Lee, Nicole L. Washington

## Abstract

**Background:** Truncating variants in *TTN* (*TTN*tvs) represent the largest known genetic cause of dilated cardiomyopathies (DCM). At the population level, even when limited to *TTN*tvs in cardiac-specific exons (hiPSI *TTN*tvs) penetrance estimates for DCM are low. Recent work shows that individuals harboring *TTN*tvs have a high prevalence of other cardiac conditions aside from heart failure, in particular, atrial fibrillation (Afib).

**Objectives:** Pinpoint the genetic footprint *TTN*-related diagnoses aside from DCM, such as Afib, and determine if vetting additional significantly-associated phenotypes better stratifies cardiomyopathy risk across *TTN* carriers.

**Methods:** We leverage longitudinal EHR and exome sequencing data from two cohorts to determine the penetrance of *TTN*tvs using multiple gene expression models against Afib, CM, and other cardiac diagnoses.

**Results:** Controlling for CM and Afib, related cardio phenotypes retain only nominal association with *TTN*tvs. An unbiased sliding window analysis of *TTN*tvs across the locus confirms the association is specific to hiPSI exons for both CM and Afib, with no meaningful associations in lowPSI exons nor improvements from LOFTEE designations. We find 34% of hiPSI *TTN*tv carriers with early Afib have a CM diagnosis – a 5-fold increase in risk over non-carriers with early Afib and 47-fold increase over population controls.

**Conclusion:** CM and Afib are often coincident in hiPSI *TTN*tv carriers, which represent varying and progressive manifestations of structurally-based heart failure. We provide statistical support for a hiPSI variant interpretation model for *TTN*tvs and evidence for the first population-level screening method with clinical utility for cardiomyopathies, especially in relation to an Afib finding.

## Introduction

Titin is a multi-use spring protein vital for cardiac contraction and relaxation, and *TTN* gene variants have long been studied in relation to dilated cardiomyopathies (DCM [MIM: 604145]). Indeed, case-control studies find rare *TTN* truncating variants (*TTN*tvs) in 15-20% of DCM cases, exceeding frequencies in matched controls(1). Recently, the ACMG upgraded *TTN* into the list of genes to screen for secondary findings in the context of clinical exome sequencing. This upgrade was based on evidence that *TTN*tvs found in the exons of heart-specific transcripts (hiPSI *TTN*tvs) pose above average risk for DCM (2–4). While there is strong evidence linking *TTN*tv to DCM, *TTN* has historically been left out of more broad population screening discussions since *TTN*tvs have variable penetrance in this context (5). Furthermore, *TTN*tvs are pervasive (roughly 1-2% of individuals) in unselected population cohorts and penetrance estimates for DCM are low, even when qualifying variants are confined to well-described hotspots, limiting the personal utility of this genetic information (3,6). A more nuanced understanding of this gene-disease relationship is needed if we are to use *TTN* genetic data in population screening to quantify future disease risk and inform clinical care.

In gene-disease relationships for which loss of function is a known mechanism of disease, it is often the case that within a definable range of the transcript, variants with similar molecular consequence confer similar disease risk. These patterns (akin to the PVS1 criterion in the ACMG variant interpretation framework), once established and validated, allow for effective phenotype-agnostic variant interpretation and make population genomic screening of rare variation for disease risk possible(7). For example, rare, loss of function variants in *LDLR*, outside of the last exon, are routinely interpreted as pathogenic for Familial Hypercholesterolemia, even if the particular variant has not been observed clinically(8,9).

*TTN* is genetically complex–a gene with 364 exons and four major structural regions (Z-disk, I-band, A-band, M-band), each with largely independent functional roles and alternative splicing both within and across these domains. Early *TTN* association studies comparing DCM cases to population variant frequencies associated DCM with truncating variants specifically in the A-band region of the protein(1). However, while odds ratios remain strongest when associations are limited to A-band variants, subsequent studies have shown that DCM risk is not exclusive to this region. The PSI (percentage spliced in) index, a calculation of exon-based cardiac expression patterns, has been leveraged to bucket *TTN* exons across all domains by their potential relevance to cardiac phenotypes(10). Population-level genetic associations remain significant when qualifying variants are defined as *TTN*tvs in constitutively expressed HiPSI exons (PSI >90)(3,4,10). Debate remains as to what extent risk extends to alternatively expressed, as opposed to constitutively expressed, cardiac exons, represented as non-hiPSI exons with intermediate PSI scores (PSI >15)(11). Establishing which regions of the *TTN* gene show statistically significant associations with relevant phenotypes, and comparing these findings to expression-based expectations, would help prioritize the genetic information that is relevant to return to patients in a genomic screening setting.

In heterogeneous population cohorts, as opposed to disease-specific cohorts, low penetrance estimates for *TTNtv* carriers may also be the result of applying a too-narrow disease phenotype to define cases. As we begin to sequence more broadly in populations, our understanding of genetic disease is changing– classic disease definitions may not completely capture population level clinical manifestations related to genetic variation at a locus(12). Relevant diagnoses may also be different depending on when in the disease course a person is assessed and first treated. For example, patients with a *TTN*tv may present later in life with DCM-related heart failure, and only including the diagnosis of DCM would falsely exclude many younger patients. By including other common symptoms such as arrhythmias and “Stage B” heart failure, it may be possible to detect the manifestations of this heart disease earlier and in a younger cohort. Indeed, *TTN*tv in hiPSI transcripts have also been associated with early onset atrial fibrillation (Afib), which may overlap with the well-established DCM association (13–15). Determining how the phenotypic footprint of *TTN*tvs extends beyond DCM may improve population-based retrospective penetrance estimates.

In this work, we leverage two population cohorts, the UK Biobank (UKB) and the Healthy Nevada Project (HNP), to evaluate the interplay between individually-significant *TTN* gene-disease associations, including DCM and Afib. We then use statistical analysis to vet variant interpretation models for *TTN*tvs across the gene and finally quantify CM risk, which together provide real world evidence to support population screening for *TTN*tvs.

## Methods

### Subjects and genetic data

We utilized the UK Biobank (UKB) plink-formatted population level exome OQFE exome files for 450k individuals (field 23155) as well as the imputed genotypes from GWAS genotyping (field 22801-22823). We also utilized 28,423 Healthy Nevada Project (HNP) samples that were sequenced and analyzed at Helix using the Exome+® assay as previously described (16). For the UKB cohort, participants range in age as of 2021 from 50 to 87 and are 55% female, while the HNP age range is from 18 to 89+ and is 68% female. The UKB is 83% British European ancestry, with another 10% of other European ancestry and 7% of other ancestries, and the HNP is 77% of general European ancestry, 14% of Hispanic ancestry, and 9% of other ancestries. No filtering was applied to the cohorts based on ancestry in the main study.

The HNP study was reviewed and approved by the University of Nevada, Reno Institutional Review Board (IRB, project 956068-12). The UKB study was approved by the North West Multicenter Research Ethics Committee, UK. All participants gave their informed, written consent prior to participation. All data used for research were deidentified.

### Translating medical records to phecodes

HNP phenotypes were processed from Epic/Clarity Electronic Health Records (EHR) data as previously described and updated as of March 2021(16). UKB data were provided from the UKB resource (accessed July 2021). For HNP, International Classification of Diseases, Ninth and Tenth Revision ICD codes and associated dates (ICD-9 and ICD-10-CM) were collected from available diagnosis tables (from problem lists, medical histories, admissions data, surgical case data, account data, claims and invoices). For UKB, ICD codes and associated dates (both ICD-9 and ICD-10) were collected from inpatient data (category 2000), cancer register (category 100092) and the first occurrences (category 1712).

We used phecodes–curated groupings of ICD codes–to reduce phenotype complexity from >20,000 ICD codes to 1,044 medically relevant phenotypes from available EHR records, specifically: ICD-9 (Phecode Map 1.2, used for both cohorts), ICD-10 (Phecode Map 1.2b to ICD-10 beta, used for UKB), and ICD-10-CM (Phecode Map 1.2b to ICD-10-CM beta, used for HNP)(17,18). When multiple ICDs from the same phecode were present for an individual, the earliest date (to year-level resolution) was used to represent that phecode. To normalize ICD dates, all dates were transformed to age at diagnosis using the difference between the ICD date and birth year of each participant.

For clinical analyses of diagnostic patterns the cohorts were filtered down to only include individuals with more than one year of diagnosis history, assessed by comparing the earliest and latest dates of any ICD code on record. These filtered cohorts included 25,493 HNP and 388,070 UKB participants.

### Annotation and PSI

Variant annotation was performed with VEP 99 (19). Coding regions were defined according to Gencode version GENCODE 33, and the Ensembl canonical transcript ENST00000589042.5 was used to determine variant consequence (20,21). However, the exon numbering we used included exon 48 from (4) which is not in the canonical transcript. Variants were restricted to CDS regions plus essential splice sites. Genotype processing for HNP data was performed in Hail 0.2.54-8526838bf99f(22).

For the collapsing analysis, samples were coded as a 1 if they had a *TTNtv* and a 0 otherwise(16). We defined *TTN*tv as LoF variants (stop_lost, start_lost, splice_donor_variant, frameshift_variant, splice_acceptor_variant, or stop_gained; however, only 1 included variant was stop_lost, and 0 were start_lost). Variants were only included if their MAF was below 0.1% in all gnomAD populations as well as locally within each population analyzed(8).

Percent spliced in (PSI) data were obtained from (4). We also annotated LoF variants as LC or HC according to LOFTEE (8).

### Power window

We developed a statistical power-based sliding window analysis technique to create a continuous pathogenicity model across the entire *TTN* locus. The basic concept of a sliding window analysis is to group variants located near each other into one unit and analyze them together to improve power, much like a gene-based collapsing analysis but at a smaller scale. Rather than size our sliding window by the number of variants or bases covered, instead our sliding window moved to maintain roughly the same number of people with a rare *TTN*tv, and thus the statistical power, within each window. When a single variant is well powered on its own, it is removed out into its own separate analysis, and the window slides past it, continuing to group surrounding variants as appropriate. We refer to this technique as a “power window” analysis.

Given the overall prevalence for cardiomyopathy (CM) and Afib of 0.6% and 5.2% in the UKB cohort with a sample size of 428,009 phenotyped individuals, any *TTN* analysis window with at least 40 carriers where the true OR is 1 has, respectively, a 99.7% (CM) and 99.5% (Afib) probability of having <3 and <7 case carriers, and thus an observed OR<12 and OR<3.5. An OR of 12 for CM and 3.5 for Afib was therefore chosen as the cutoff for a window to be considered associated with the trait. These OR cutoffs are also consistent with the overall OR observed for hiPSI exons in this analysis more generally (CM: 12.2, 95%CI 10.2-14.5; Afib: 3.3, 95%CI 2.9-3.8). With 40 variant carriers within each window, the power for discovery is the same for each window as our analysis slides across the gene. Bases that fall between the boundaries of an associated window are assigned the same value in the testing set as that window has in the training set. Thus, new mutations that do not occur in the original dataset can still be assigned a value based on their location in the testing set. This also means that exons with no variation in the training set are assigned a value based on the values of their surrounding exons, which defines whether or not they are within the boundaries of an associated window. Our model tested 1,336 windows. Each window includes a mean of 20.5 variants (median 22, range 1-35) and 7.2 exons (median 7, range 1-22). Variants are a mean of 100.3 coding bases apart (median 63, range 0-759).

### Statistical analysis

We used regenie for genetic analyses as previously described (16,23,24). Our analysis in regenie builds a whole genome regression model using common variants to account for the effects of relatedness and population stratification, and it accounts for situations where there is an extreme case-control imbalance, which can lead to test statistic inflation with other analysis methods. The covariates we included were age, sex, age*sex, age*age, sex*age*age, and bioinformatics pipeline version as appropriate. As previously described, a representative set of 184,445 coding and noncoding LD-pruned, high-quality common variants were identified for building the whole genome regression model.

We analyzed all ancestries together because when collapsing rare (MAF<0.1%) causal variants across a gene and analyzing with a linear mixed model or whole genome regression, signals tend to be consistent whether restricting to one ancestry or analyzing across all ancestires. This method works in this setting because analyses of collapsed rare variants are less influenced by ethnic background than are analyses of the common variants used in a typical GWAS, in large part because causal variants are being grouped together as opposed to tagging variants. **Table S1** breaks down the hiPSI *TTN*tv counts by ancestry.

Other statistical analyses were run using the statsmodel package in python 3.7.7. For binary variables, logistic regression was used; for quantitative variables, linear regression was used after rank-based inverse normal transformation.

### Homozygotes and compound heterozygotes

As a further characterization of regions of *TTN* containing pathogenic variants, we looked at individuals who were homozygous for a *TTN*tv or were heterozygous for more than one *TTN*tv. In the UKB450K, we identified 16 individuals who were homozygous for a *TTN*tv, all of which were in loPSI exons. We also identified 105 individuals who were heterozygous for two *TTN*tvs. Of these, 83 were unlikely to be true compound heterozygotes as the two variants were within 10 bp of each other or seen together in multiple individuals (although 15kb apart, chr2:178661989:GTTTTC:G was observed in combination with chr2:178677634:TG:T all 7 times that it was seen in the UKB450K; and although 120kb apart, chr2:178534380:T:A was observed with chr2:178653262:T:A both times it was seen in the UKB450K: all of these variants are in loPSI exons). The remaining potential compound heterozygotes had their two variants at least 4kb apart from each other, and their specific combination of variants was seen only in one individual in the UKB450K. Of these, in 9 individuals both variants were in loPSI exons, and in 13 individuals one variant was in a loPSI exon and one in a hiPSI exon. No individuals with 2 hiPSI *TTN*tvs were observed, despite hiPSI carriers constituting 34% of those with *TTN*tvs.

## Results

### *TTN*tv associations with cardio phenotypes are driven by atrial fibrillation and cardiomyopathy

Our previous work examined rare LoF variants in *TTN* (equivalent to and hereafter *TTN*tvs) with a gene-based collapsing approach in two population-based cohorts (∼25k individuals from the Healthy Nevada Project (HNP) and ∼200K from the UKB). Through these analyses we identified genome-wide significant associations with seven heart phenotypes (“phecodes”, see methods): primary/intrinsic cardiomyopathies (CM), heart failure not otherwise specified (NOS), atrial fibrillation and flutter (Afib), congestive heart failure (CHF) NOS, nonrheumatic mitral valve disorders, mitral valve disease, and tachycardia NOS(24). While DCM and Afib are the most well-described population-level associations, *TTN*tvs have been implicated in a spectrum of heart disease and subclinical functional measures, including each of the phenotypes identified in these analyses (13,14,25–27). Here, the association we identify with DCM also includes some other less common CM categories (phecode 425.1 includes not only ICD10 code I42.0, cardiomyopathy, but also I42.3, endomyocardial (eosinophilic) disease (n=2 cases), I42.4, endocardial fibroelastosis; I42.5, other restrictive cardiomyopathy; I42.8, other cardiomyopathies, and I42.9, unspecified cardiomyopathy). We include this broader CM phenotype as opposed to only DCM because there was an enrichment for *TTN*tv carriers with this aggregate phenotype even after removing the strictly DCM cases (OR=3.3). Similar to previous reports, the penetrance estimates for each discrete phenotype was low (2-9%), limiting the clinical utility of each individual result for carriers. While each phenotypic association may represent a discrete disease, we wanted to test if instead, the multiple associations reflect varying and compounding outcomes that result from underlying structural, functional, and metabolic changes stemming from *TTN*tvs (3,13,15,28–30).

To test this hypothesis, we leveraged a larger dataset of ∼430,000 clinico-genomic records from the UK Biobank (UKB450K). We found that 6 of the 7 previously associated individual phenotypes maintained genome-wide significant p-values (p<5-e8) from this updated gene-based collapsing analysis, and the p-value for tachycardia NOS was no longer significant (p<1e-5) (Figure 1). The highest OR was found for CM (OR=5.5, p-value< 3.5e-63), with the other ORs lying within the range of 1.8-2.2. Given that CM and Afib are already firmly established as associated with *TTN*tv(3,14), we next conditioned on these two diagnoses to determine whether their associations were at least partially independent and whether other phenotypes had independent associations as well. We found that the association between CM and *TTN*tv was still statistically significant after controlling for Afib (p=1.9e-52), and the association between Afib and *TTN*tv was also still statistically significant after controlling for CM (p=4.2e-18). However, after controlling for both of these conditions, none of the other phenotypes maintained genome-wide significance. The best remaining association was with heart failure NOS (p=1e-5; other phenotype p-values ranged from 0.02-0.002).

**Figure 1.**
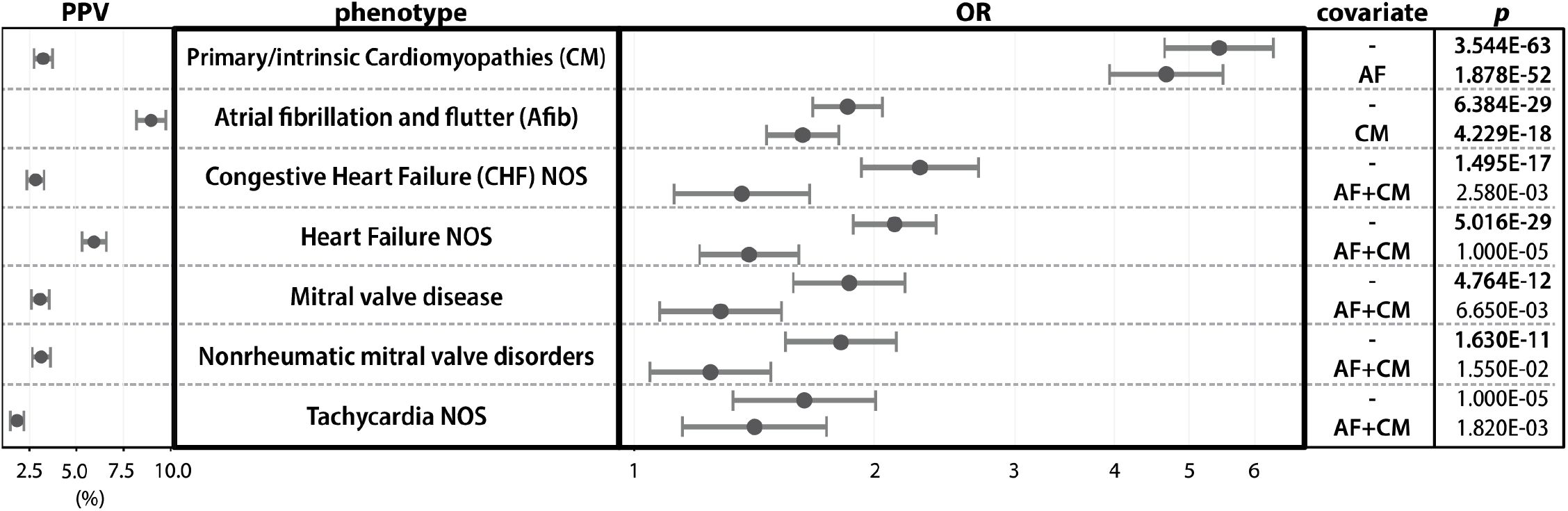
*TTN*tv genetic association analyses. Plotted is the odds ratio (OR) and positive predictive value (PPV) for the 7 phenotypes identified as significantly associated with *TTN*tvs in our prior studies (24). The values shown and corresponding p-values are updated based on analysis of the 450k UKB exomes.

Overall, these controlled analyses show that Afib and CM represent independent but overlapping associations with *TTN*tvs and support the hypothesis that these diagnoses may share underlying pathophysiology, such as impared sarcomere function, in *TTN*tv carriers(13,14). Additional phenotypic associations (ie mitral valve, heart failure, and tachycardia) can be largely explained by controlling for Afib and CM but because they are still enriched in *TTN*tv carriers may represent complications not traditionally considered in relation to these core diagnoses. Future studies of *TTN*tv carriers may benefit from broad cardiac phenotyping to further our understanding of these trends. For the remainder of this study, we will focus our analysis on the gene-disease relationship between CM, Afib and *TTN*tvs in unselected population cohorts.

### An unbiased sliding window statistical analysis confirms hiPSI variant prioritization for both Afib and CM risk

With confirmation of each association at the gene level, we next determined if elevated Afib and CM risk localized to truncating variants in similar or varying regions of the *TTN* locus. Given previous work on DCM, we expected the signal would likely be localized to constitutively expressed cardiac exons (hiPSI) and that variants in those exons would have a higher penetrance (**Figure 2A**). However, the hiPSI exon definition varies slightly based on the expression dataset used to determine PSI values, with models derived from both the GTEx dataset and from left ventricular tissue from DCM patients available (3,4). Further, while debate remains around the mechanism of disease of *TTNtv*s in cardiac transcripts, there is evidence for haploinsufficiency, and nonsense-mediated decay considerations (i.e., LOFTEE) may improve *TTNtv* selection(8,31).

**Figure 2.**
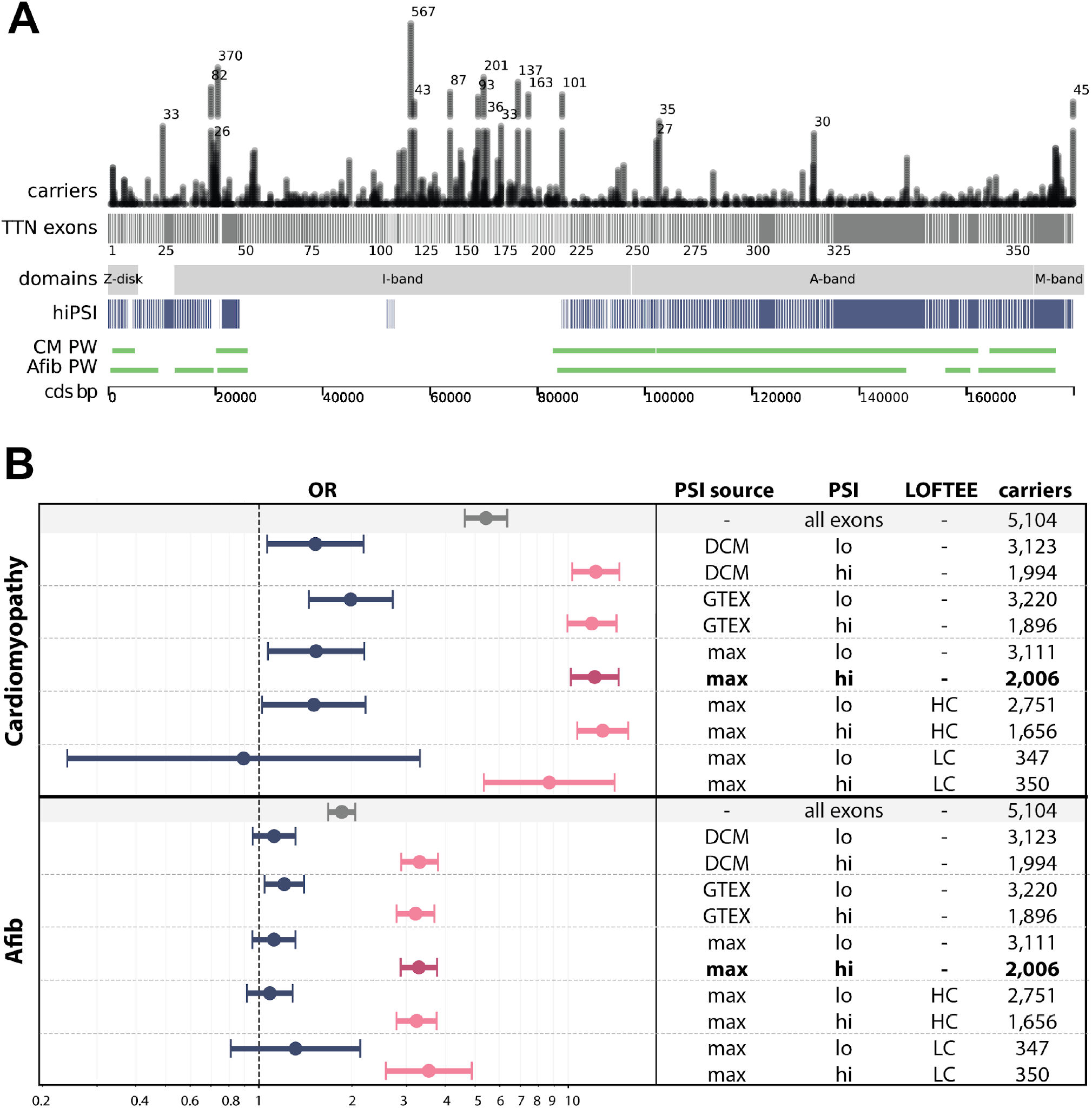
*TTN* variants and regions that are associated with CM and Afib. **(A)** TTN*tv carriers*: shown above the *TTN* canonical exon track, each dot represents an individual carrier and are stacked at a given position. For brevity, carriers are trimmed to 35 and total number of carriers is indicated when >40. Carriers are from the UKB450K cohort. TTN *exons*: canonical exons (gray; introns not to scale), with the exon number indicated below track. *Domains*: major protein bands (Z-disk, I-band, A-band, M-band)(4). *cardio hiPSI*: exons with “percent spliced in” > 90 per exon, according to the max of either GTEx or DCM patients for heart tissue (blue). *PW*: merger of all power windows with an OR outside the 99% confidence bound for a true OR of 1 when sampling 40 rare-variant carriers. Coding position indicated. **(B)** Association between *TTN*tvs and CM (cardiomyopathy) or Afib (atrial fibrillation). The whole gene is shown in gray, and the hiPSI exons (PSI>90) are shown in pink, while the loPSI exons (PSI<=90) are shown in blue. This was performed separately for PSI based on DCM patients, GTEX (GTEx) data, or the maximum of those too. The variants are also broken up by whether LOFTEE categorized them as HC (high confidence) or LC (low confidence).

To this end, we leveraged the clinical data to identify the regions of the gene where there was statistical evidence to support an association between *TTN*tvs and either CM or Afib. We identified these regions using a sliding window strategy to maintain roughly the same number of people with a rare *TTN*tv, and thus the statistical power, within each window (see Methods). Using this “power window” technique, an analysis of CM and Afib in the UKB450K cohort identified nearly all the hiPSI regions of the gene and almost no loPSI regions (**Figure 2A**). Specifically, in and near the Z band (exons 2-25, all hiPSI except 11 and 12), part of the I band (exons 29-53 and 217-252, all hiPSI except exons 217-219, 225, and 243), the A band (exons 253-358, all hiPSI), and part of the M band (part of hiPSI exon 359) all showed statistical enrichment for CM or Afib among *TTN*tv carriers (**Table S2**). This association was found despite the statistical analysis being blind to the PSI levels of each exon. This result demonstrates that the signal for *TTNt*vs with both CM and Afib is driven by variants in the hiPSI exons across all major protein domains, matching the previously observed association between DCM and hiPSI *TTN*tvs(3).

We also considered if exons with PSI values indicative of alternative expression (PSI between 15 and 90) were enriched for CM or Afib phenotypes using this sliding window method. We found that 77% and 90% of PSI>90 exons overlapped a window with a significant association with CM and Afib phenotypes, respectively, whereas these numbers dropped to 3%-15% of exons in other PSI ranges for these phenotypes (**Figure S1**). These results confirm that a hiPSI cutoff of >90 is appropriate for exon selection for both CM and Afib. Furthermore, we identified 38 individuals in the UKB450K who were homozygous or compound heterozygous for a *TTN*tv: none of these individuals had two *TTN*tvs in exons with PSI>90, supporting the specificity of this hiPSI model (see supplement for details).

Next, we assessed whether hiPSI models calculated from expression data from the left ventricle of patients with DCM or from the heart tissue of GTEx participants were more predictive of CM or Afib at the population level. Using the now confirmed PSI >90 cutoff, we found that there was little difference in the magnitude and separation of odds ratios for hiPSI groupings stemming from each model (**Figure 2B**). With support for hiPSI models that are derived from either expression data source, we decided to use the maximum PSI value between these two sources for each exon to build a hiPSI model for population screening. Finally, we assessed the utility of using LOFTEE in addition to a hiPSI exon filter in distinguishing pathogenic from benign *TTN*tvs. We found that LOFTEE annotation did not meaningfully distinguish pathogenic *TTN*tvs from benign ones, and that hiPSI designation was the most important factor for pathogenicity (**Figure 2B**).

Given these results, we performed our remaining analyses on all exons with >90% maximum PSI (hiPSI), disregarding LOFTEE status. Restricting to hiPSI exons removed 34% of the *TTN*tvs, assigning them to a likely benign status based on their exon location, and reclassified 61% of the individuals with *TTN*tvs from carriers to non-carriers. As expected, this refinement substantially improved both the p-value (from p=3.5e-63 (CM) and 6.4e-29 (Afib) for the entire gene to p=1.1e-93 and 1.3e-52) and the PPV (from 3% and 9% for the entire gene to 7% and 14%) (**Figure 2**, <u>Table S1</u>).

### Thirty-four percent of hiPSI *TTN*tv carriers with early Afib have a CM diagnosis

While the penetrance for CM as an individual diagnosis, even when limiting to hiPSI *TTN*tv carriers, remains below 10% in the UKB450K, our genetic analyses suggest the more prevalent Afib diagnosis (penetrance 14%) may also be relevant to consider. To that end, we next investigated if accounting for Afib would identify a subset of hiPSI *TTN*tv individuals at highest risk for CM. (Basic cohort demographics and rates for CM and Afib phecodes in **Table S3**).

We find that across all individuals with an Afib diagnosis, the prevalence of a CM diagnosis varies by *TTN* genotype, with the highest risk residing amongst hiPSI *TTN*tv carriers with an Afib diagnosis before age 60 (“early Afib”)(13,14). Specifically, CM diagnoses in individuals with early Afib are recorded in 7% of non-carriers and 5% of loPSI *TTNtv* carriers as compared to 34% of individuals with hiPSI *TTN*tvs — an over five-fold increase in CM risk by genotype status **(Figure 3**, p=4.8e-56 after controlling for age and sex). These trends replicate in the HNP cohort (**Figure S2**). To determine if this link between diagnoses is indicative of a disease progression, we looked more specifically at the temporal patterning of diagnoses in *TTN*tv hiPSI carriers with both Afib and CM. Looking at year-level resolution, we find that Afib either predates or is concurrently diagnosed with CM in 72% of individuals with both diagnoses (**Table S3**). Additionally, we observe 87% of hiPSI *TTN*tv carriers with Afib and CM diagnoses also carry a diagnosis code for heart failure, as compared to 70-75% for non-carriers and loPSI TTNtv carriers with Afib and CM diagnoses. Taken together, our retrospective analysis of EHR diagnosis data supports a link between Afib and CM with likely progression to heart failure in hiPSI *TTN*tv carriers, especially when Afib is diagnosed before age 60.

**Figure 3.**
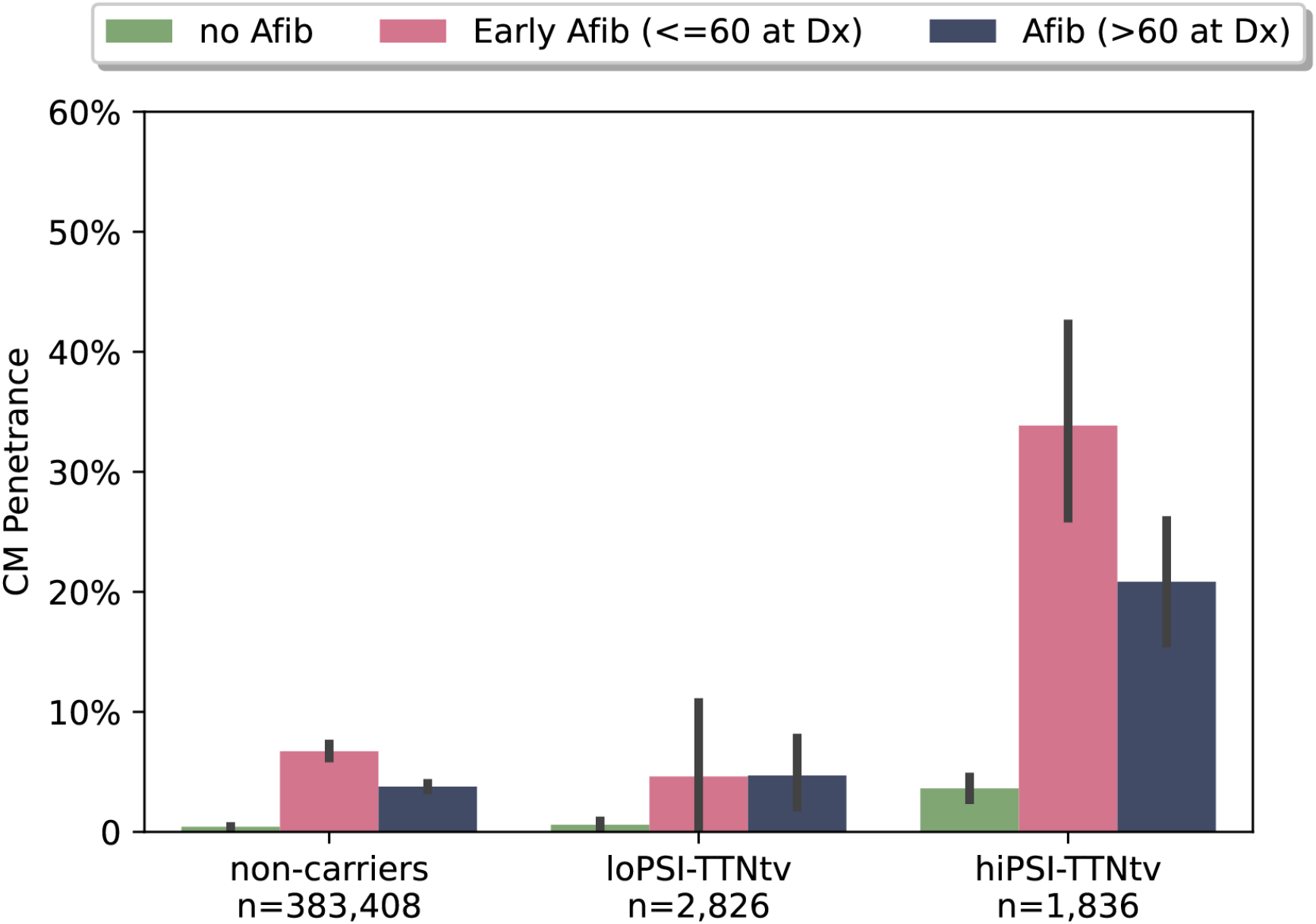
CM Prevalence in UKB participants stratified by *TTN*tv status and Afib diagnosis. All UKB450K participants with ICD codes spanning more than one year in available EHR records (n=388,070) were separated by their *TTN* genotype status (non-carriers, LoPSI, and HiPSI carriers) and by the presence and, where relevant, onset of Afib. The penetrance and bootstrapped 95% confidence interval for CM is plotted on the y-axis for each of these groupings. While those with an early Afib diagnosis (pink bars) show the highest prevalence of CM, hiPSI *TTN*tvs are at more than five-fold greater risk for CM than any other group (p=4.8e-56 after controlling for age and sex).

Given the strong relationship between these diagnoses, evidence of shared genetic etiology, and patterning in available records, we hypothesize that it may be reasonable to screen for Afib in individuals with a qualifying *TTN*tv, which may represent an early sign of a structurally-based heart failure. Finally, we also observe a small but significant difference in age of onset of both Afib and CM between hiPSI carriers and non-carriers (CM median age 61 vs 65 and Afib median age 65 vs 68 in hiPSI and non-carriers, respectively, p=0.0004 and p=4.7e-11, **Figure S3**). This shift in overall onset of both diagnoses suggests that screening hiPSI *TTN*tv carriers for signs of heart failure such as Afib earlier in the life course may have clinical value.

## Discussion

The link between *TTN* and DCM is well known, yet due to low penetrance estimates, *TTN* has not been suitable for general population screening. It is well-known that individuals often carry a range of structurally-related cardio diagnoses and that penetrance estimates across *TTN*tv-related diagnoses improve when carriers are limited to only those with truncating variants in exons with constitutive expression in heart tissue (hiPSI exons). In this work, we focus on understanding the interplay between the various cardio diagnoses associated with *TTN*tvs and the underlying genetics for each association. We find that significant, population-level genetic associations remain for both CM and Afib phenotypes when controlling for the other, while all other associations can be largely explained by these two phenotypes. Using a statistical-based analysis across the locus we show that a hiPSI-based variant interpretation model faithfully represents both the CM and Afib associations and, in line with other reports, penetrance increases for both phenotypes (9% for CM and 14% for Afib) after subsetting to only those with hiPSI *TTN*tvs. Finally, to understand the connection between these diagnoses we perform a retrospective analysis of these diagnoses in individuals with and without hiPSI *TTN*tv. We find that when focusing on individuals with an early Afib diagnosis, 34% of hiPSI *TTN*tv carriers also carry a diagnosis of CM, over a five-fold enrichment compared to noncarriers with early Afib and a 47-fold increase in CM risk compared to the general population.

### Strategy for population screening for hiPSI *TTN*tvs

There are currently no screening recommendations for genetic cardiomyopathies outside known family history. The high prevalence of CM in hiPSI *TTN*tv carriers with early Afib suggests that population screening for hiPSI *TTN*tvs (∼0.5% of a cohort) and monitoring for Afib in these individuals may be one strategy to identify those at high risk of CM earlier in the disease course. Since early detection enables early intervention, and since *TTN*tv carriers with DCM are known to respond well to standard of care interventions(27), *TTN*tv screening in conjunction with routine monitoring may be an effective strategy to improve outcomes and to reduce the incidence of heart failure in the population(31,32). Supporting this screening strategy, we find that Afib often predates or is at least concurrently diagnosed with CM in hiPSI *TTN*tv carriers.

### Atrial fibrillation and wearables

As wearables increase in popularity and function, awareness of atrial fibrillation is likely to increase. Understanding heart disease risk in relation to an Afib finding from the wearable device will likely become a more common medical inquiry (33). These findings can be hard to interpret as an individual data point in a patient, as they could represent one-off occurrences as opposed to a sign of elevated cardiovascular disease risk. Based on our findings surrounding clinically-diagnosed Afib, CM, and *TTN*tvs, further studies comparing the relationship between Afib diagnoses collected via wearables and clinical-based assessments in hiPSI *TTN*tv carriers should be performed.

### Limitations

One of the major limitations of this study (and most studies that rely on retrospective analysis of the EHR) is the lack of detailed phenotyping, especially of lower risk phenotypes, over a longitudinal timespan in the participants. Despite EHRs’ comprehensive datasets, many data points are not recorded with the desired frequency, coding patterns vary by both physician and health system, and initiation dates of events are suboptimal. Thus, penetrance estimates will likely be higher with more detailed evaluations. A lack of a standard screening strategy may at least partially explain the often concurrent documentation of Afib and CM diagnoses in our cohorts, a trend which was also reported in a recent epidemiological study of all cardiomyopathies(28). For all phenotypes, penetrance and expressivity may also depend on environmental stressors including alcohol, chemotherapy, and pregnancy, which can put extra stress on the heart and could precipitate otherwise subclinical disease in some *TTN*tv carriers (34). We were encouraged by the replication of the Afib and CM coding trends across two healthy system cohorts with different diagnosis rates (UKB and HNP, **Table S3**), but follow up studies with more detailed and longitudinal phenotyping should be completed to confirm these retrospective trends. Finally, while our analysis showed similar trends between different ethnicities (**Table S1**), the limited sample sizes for non-European ancestries makes it difficult to show if there are ancestry-specific differences in the effects of variants in this gene, as previously suggested(3).

Our analysis with the power window technique showcases the power of using clinico-genomic data, in the spirit of a traditional gene knockout experiment, to improve variant interpretation and better understand the genetic mechanism of disease at a locus. The I-band in particular appears to be a hotspot for *TTN*tvs as compared to the rest of the gene but without a proportional increase in cases, consistent with most of this region being spliced out in cardiac-specific transcripts. Although our method provided statistical confirmation for the significance of hiPSI *TTN* exons in general, we did not find strong statistical support for hiPSI exons 26-28, 108-113, part of 359, or 360-364 to be associated with CM or Afib. The inclusion of those exons may be reconsidered if further studies confirm their lack of association. In particular, the variants in the C-terminal M-band exons showed no association with the phenotypes, consistent with truncating mutations at the ends of genes often having benign effects. The other lack of associations for the other exons are more complicated: exons 26-28 were close to the significance threshold, and exons 108-113 of the I band were an isolated set of hiPSI exons that only contained 13 *TTN*tv carriers in this dataset and thus were not well powered for analysis. However, the surrounding exons from 91-118 had PSI levels of 80+ and 216 *TTN*tv carriers and still did not indicate a strong association in this region. In summary, although 1.17% of individuals in the general population appear to harbor a *TTN*tv, in reality the incidence drops to 0.5% based on hiPSI screening regions, representing only 56% of the coding portion of the gene.

## Conclusion

Through the analysis of paired clinical and exome data from two population cohorts, we demonstrate a path forward for *TTN* population screening. We provide statistical confirmation that *TTN*tvs in hiPSI exons are associated with CM and Afib in population cohorts, and based on a retrospective analysis of diagnosis show that basic screening measures to catch Afib, which is likely undiagnosed at onset, may help further prioritize those at high risk.

## Data Availability

Statistics relating to the CM and Afib power window analyses for all TTN exons, calculated using the UKB450K are available in Table S2. UKB data are available for download (https://www.ukbiobank.ac.uk/) to qualified researchers. The HNP data are available to qualified researchers upon reasonable request and with permission of the Institute for Health Innovation (IHI) and Helix. Researchers who would like to obtain the raw genotype data related to this study will be presented with a data user agreement which requires that no participants will be re-identified and no data will be shared between individuals or uploaded onto public domains. The IHI encourages and collaborates with scientific researchers on an individual basis. Examples of restrictions that will be considered in requests to data access include but are not limited to (1) whether the request comes from an academic institution in good standing and will collaborate with our team to protect the privacy of the participants and the security of the data requested, (2) type and amount of data requested, (3) feasibility of the research suggested, (4) amount of resource allocation for the IHI and Renown Hospital required to support the collaboration. Any correspondence and data availability requests related to HNP should be addressed to JG at or Craig Kugler.

## Abbreviations

PPV: positive predictive value
*TTN*tvs: *TTN* truncating variants
LoF: loss of function
hiPSI: percent spliced in >90%
loPSI: percent spliced in <=90%
DCM-dilated cardiomyopathies CM: cardiomyopathy
Afib: atrial fibrillation
PVS1: pathogenic very strong criteria 1 from the ACMG variant interpretation rubric

## Acknowledgements

This research has been conducted using the UK Biobank Resource under Application Number 40436. We acknowledge the entire Helix Bioinformatics team for their contributions to the production exome sequencing pipeline. We thank all of the genomic representatives of the Healthy Nevada Project (HNP). We thank Renown Health and DRI marketing for helping to launch the HNP.

## Data code and availability

Statistics relating to the CM and Afib power window analyses for all *TTN* exons, calculated using the UKB450K are available in Table S2. UKB data are available for download (https://www.ukbiobank.ac.uk/) to qualified researchers. The HNP data are available to qualified researchers upon reasonable request and with permission of the Institute for Health Innovation (IHI) and Helix. Researchers who would like to obtain the raw genotype data related to this study will be presented with a data user agreement which requires that no participants will be re-identified and no data will be shared between individuals or uploaded onto public domains. The IHI encourages and collaborates with scientific researchers on an individual basis. Examples of restrictions that will be considered in requests to data access include but are not limited to (1) whether the request comes from an academic institution in good standing and will collaborate with our team to protect the privacy of the participants and the security of the data requested, (2) type and amount of data requested, (3) feasibility of the research suggested, (4) amount of resource allocation for the IHI and Renown Hospital required to support the collaboration. Any correspondence and data availability requests related to HNP should be addressed to JG at or Craig Kugler.

## Supplemental Figures

**Fig S1.**
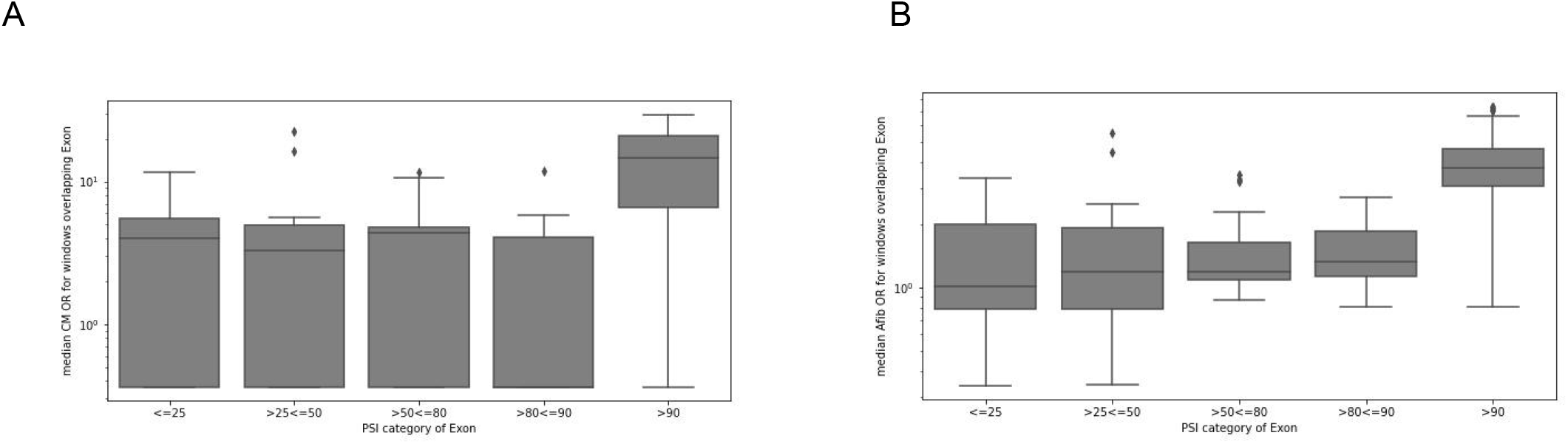
Median odds ratio (OR) of power windows overlapping each exon, split up into PSI categories. A) Cardiomyopathy (CM); B) Atrial fibrillation (Afib). The PSI value is the max PSI value per exon, from DCM patients or GTEx. There is no support for exons with PSI <=90 to be associated with CM or Afib. Note nearly all exons in the PSI>90 category are PSI 100, making it difficult to tell where the cutoff for association is within the >90 range.

**Fig S2.**
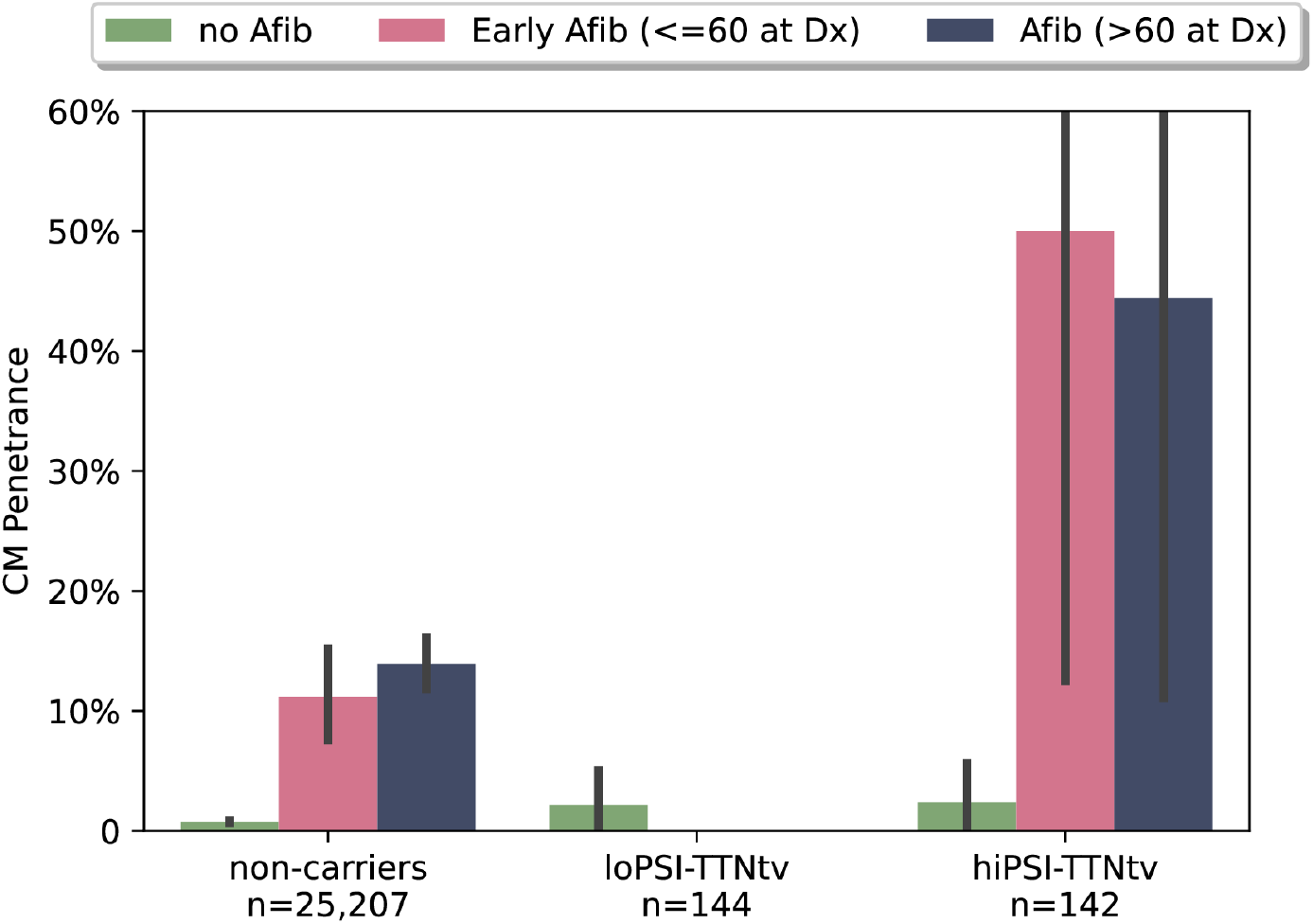
CM Prevalence in HNP participants stratified by *TTN*tv status and Afib diagnosis. All HNP25K participants with ICD codes spanning more than one year in available EHR records (n=25,493) were separated by their *TTN* genotype status (non-carriers, LoPSI, and HiPSI carriers as described Results). Each genotypic group was further separated by the presence and, where relevant, onset of an Afib phecode. The penetrance of CM phecode is plotted on the y-axis for each of these groupings, with a bootstrapped 95% confidence interval indicated for each estimate. While those with an early Afib diagnosis (pink bars) show the highest penetrance for CM across each genotypic subset, hiPSI *TTN*tvs are at more than five fold greater risk for CM than any other group (p=0.0003 after controlling for age and sex)

**Fig S3.**
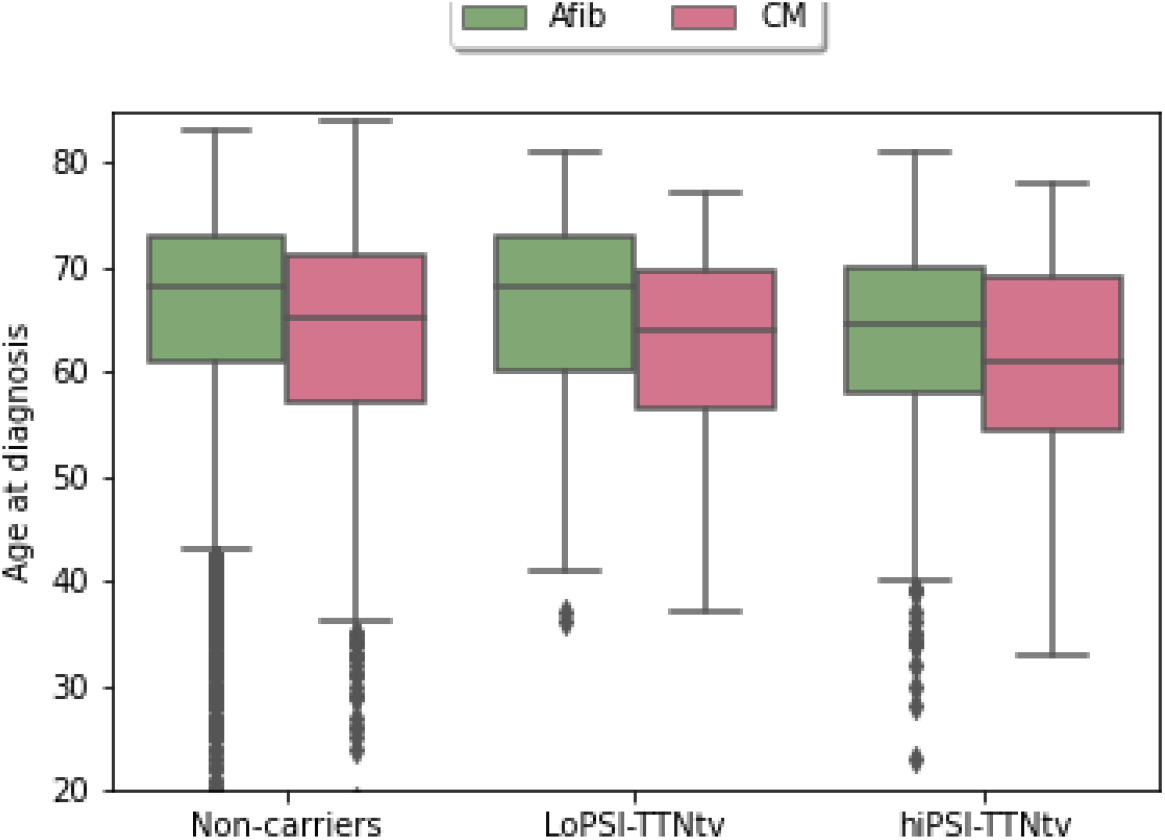
Age of onset for Afib and CM diagnoses by *TTN*tv status in the UKB450K. All UKB participants with at least one year of ICD history (n=388,070) were separated by their *TTN* genotype status (non-carriers, LoPSI, and HiPSI carriers as described Results). Plotted is the distribution of age at diagnosis for those with either an Afib (pink) or CM (green) phecode in available medical records. Lines represent mean, boxes and whiskers represent quartiles in either direction. Outliers are plotted as individual points. Age of onset for both Afib and CM is lower in hiPSI-*TTN*tvs compared to non-carriers (CM median age 61 vs 65 and Afib median age 65 vs 68 in hiPSI and non-carriers, respectively, p=0.0004 and p=4.7e-11)

