## Supplemental Tables for "*TTN* truncating variants in hiPSI exons show high penetrance for cardiomyopathy in carriers with atrial fibrillation"

| Table | Description |
|---|---|
| S1 | Association stats for CM and Afib with TTN rare LoF in PSI>90 exons across different ancestries. |
| S2 | TTN exons and metadata |
| S3 | UKB and HNP cohort demographics and phecode summary |
| S4 | Breakdown of temprotal diagnosis patterning of individuals with both atrial fibrillation (Afib) and cardiomyopathy (CM) phecodes in the UKB450k |

**Table S1. Association stats for CM and Afib with TTN rare LoF in PSI>90 exons across different ancestries.**

| Phenotype | Ancestry | UKB n | UKB OR (model with covariates) | UKB PPV | UKB phenotype frequency | UKB percent carriers | UKB n observed case carriers | UKB n expected case carriers if OR=1 (no association) | UKB n expected case carriers if OR=12 (CM) or 3.5 (Afib) | HNP n | HNP OR (model with covariates) | HNP PPV | HNP phenotype frequency | HNP percent carriers | HNP n observed case carriers | HNP n expected case carriers if OR=1 (no association) | HNP n expected case carriers if OR=12 (CM) or 3.5 (Afib) |
|---|---|---|---|---|---|---|---|---|---|---|---|---|---|---|---|---|---|
| CM | African | 7372 | 11.5739 | 7.69% | 0.95% | 0.53% | 3 | 0 | 4 | 547 | 0.334678 | 0.00% | 2.38% | 0.55% | 0 | 0 | 1 |
| CM | East Asian | 2803 | 0.362486 | 0.00% | 0.36% | 0.50% | 0 | 0 | 1 | 867 | 20.30952 | 14.29% | 0.93% | 0.81% | 1 | 0 | 1 |
| CM | South Asian | 6715 | 9.90187 | 4.17% | 0.57% | 0.71% | 2 | 0 | 3 | 167 | NA | 0.00% | 0.00% | 0.60% | 0 | 0 | 0 |
| CM | Hispanic | NA | NA | NA | NA | NA | NA | NA | NA | 4054 | 0.361843 | 0.00% | 0.27% | 0.54% | 0 | 0 | 1 |
| CM | British | 359867 | 12.1767 | 6.72% | 0.62% | 0.47% | 113 | 10 | 113 | NA | NA | NA | NA | NA | NA | NA | NA |
| CM | Other European | 43897 | 18.9206 | 9.42% | 0.62% | 0.44% | 18 | 1 | 13 | 21795 | 7.75171 | 7.38% | 1.40% | 0.56% | 9 | 2 | 19 |
| CM | Other | 7355 | 8.09018 | 3.13% | 0.77% | 0.44% | 1 | 0 | 3 | 993 | 0.313273 | 0.00% | 0.70% | 0.40% | 0 | 0 | 0 |
| Afib | African | 7372 | 1.24925 | 2.56% | 2.29% | 0.53% | 1 | 1 | 3 | 547 | 0.266907 | 0.00% | 2.56% | 0.55% | 0 | 0 | 1 |
| Afib | East Asian | 2803 | 0.358128 | 0.00% | 2.35% | 0.50% | 0 | 0 | 1 | 867 | 80.3492 | 14.29% | 1.50% | 0.81% | 1 | 0 | 0 |
| Afib | South Asian | 6715 | 2.36105 | 6.25% | 2.71% | 0.71% | 3 | 1 | 4 | 167 | NA | 0.00% | 3.09% | 0.60% | 0 | 0 | 0 |
| Afib | Hispanic | NA | NA | NA | NA | NA | NA | NA | NA | 4054 | 0.320162 | 0.00% | 1.38% | 0.54% | 0 | 0 | 1 |
| Afib | British | 359867 | 3.42253 | 14.33% | 5.42% | 0.47% | 241 | 83 | 287 | NA | NA | NA | NA | NA | NA | NA | NA |
| Afib | Other European | 43897 | 3.27357 | 14.14% | 4.64% | 0.44% | 27 | 8 | 28 | 21795 | 3.18804 | 13.11% | 5.12% | 0.56% | 16 | 6 | 20 |
| Afib | Other | 7355 | 0.816784 | 3.13% | 3.66% | 0.44% | 1 | 1 | 4 | 993 | 0.365234 | 0.00% | 2.01% | 0.40% | 0 | 0 | 0 |

**Table S2 - TTN exons and metadata**

| Exon | hg38 start | hg38 end | band | Coding length | GTEx PSI | DCM PSI | Max PSI | CM: min OR for overlapping windows | CM: max OR for overlapping windows | CM: median OR for overlapping windows | Afib: min OR for overlapping windows | Afib: max OR for overlapping windows | Afib: median OR for overlapping windows | n windows | CM: n sig windows | Afib: n sig windows | Note |
|---|---|---|---|---|---|---|---|---|---|---|---|---|---|---|---|---|---|
| Exon_1 | 178807212 | 178807423 | Z-disk | 0 | 100 | 100 | 100 | NA | NA | NA | NA | NA | NA | NA | NA | NA | |
| Exon_2 | 178804552 | 178804655 | Z-disk | 91 | 100 | 100 | 100 | 10.2011 | 10.2011 | 10.2011 | 5.43602 | 5.43602 | 5.43602 | 1 | 0 | 1 | |
| Exon_3 | 178802138 | 178802341 | Z-disk | 204 | 100 | 100 | 100 | 5.07643 | 26.0581 | 7.78375 | 3.06521 | 6.54425 | 3.76166 | 13 | 1 | 9 | |
| Exon_4 | 178800395 | 178800682 | Z-disk | 288 | 100 | 100 | 100 | 5.07643 | 26.0581 | 9.50381 | 3.06521 | 6.83489 | 3.77572 | 18 | 2 | 14 | |
| Exon_5 | 178799825 | 178799910 | Z-disk | 86 | 100 | 100 | 100 | 5.07643 | 26.0581 | 9.74451 | 3.06521 | 6.83489 | 3.77572 | 20 | 4 | 16 | |
| Exon_6 | 178799487 | 178799731 | Z-disk | 245 | 100 | 100 | 100 | 5.07643 | 26.0581 | 9.74451 | 3.06521 | 6.83489 | 3.77572 | 21 | 5 | 17 | |
| Exon_7 | 178794922 | 178795252 | Z-disk | 331 | 100 | 100 | 100 | 0.364685 | 26.0581 | 9.50381 | 3.06521 | 6.83489 | 3.76869 | 24 | 5 | 19 | |
| Exon_8 | 178794399 | 178794551 | Z-disk | 153 | 100 | 100 | 100 | 0.36444 | 14.4402 | 6.327255 | 2.40663 | 6.83489 | 3.572425 | 30 | 4 | 17 | |
| Exon_9 | 178793404 | 178793541 | Z-disk | 138 | 100 | 100 | 100 | 0.36444 | 14.4402 | 0.36488 | 2.40663 | 6.83489 | 3.331245 | 36 | 4 | 16 | |
| Exon_10 | 178792072 | 178792197 | Z-disk | 126 | 100 | 100 | 100 | 0.36444 | 14.4402 | 0.36488 | 2.40663 | 6.83489 | 3.38666 | 36 | 4 | 17 | |
| Exon_11 | 178790708 | 178790845 | Z-disk | 138 | 63 | 51 | 63 | 0.36444 | 14.4402 | 0.364866 | 2.40663 | 6.83489 | 3.2314 | 35 | 4 | 17 | |
| Exon_12 | 178789978 | 178790115 | Z-disk | 138 | 75 | 79 | 79 | 0.36444 | 14.4402 | 0.364685 | 2.40663 | 6.83489 | 3.2314 | 33 | 4 | 15 | |
| Exon_13 | 178789360 | 178789497 | Z-disk | 138 | 95 | 96 | 96 | 0.36444 | 14.4402 | 0.3646435 | 2.40663 | 4.23679 | 3.121685 | 28 | 3 | 10 | |
| Exon_14 | 178785848 | 178786141 | Z-disk | 294 | 100 | 100 | 100 | 0.36444 | 14.4402 | 0.364638 | 2.40663 | 4.23679 | 3.09685 | 27 | 2 | 9 | |
| Exon_15 | 178785620 | 178785742 | Z-disk | 123 | 99 | 99 | 99 | 0.36444 | 6.80855 | 0.364576 | 2.40663 | 4.23679 | 3.07127 | 21 | 0 | 5 | |
| Exon_16 | 178784070 | 178784351 | near Z-disk | 282 | 100 | 99 | 100 | 0.36444 | 6.80855 | 0.364576 | 2.40663 | 4.23679 | 3.07127 | 21 | 0 | 5 | |
| Exon_17 | 178783720 | 178783785 | near Z-disk | 66 | 100 | 99 | 100 | 0.364462 | 6.80855 | 0.364577 | 2.58605 | 4.23679 | 3.120565 | 18 | 0 | 5 | |
| Exon_18 | 178782806 | 178783064 | near Z-disk | 259 | 100 | 100 | 100 | 0.364462 | 6.80855 | 0.364578 | 2.58605 | 4.23679 | 3.062405 | 20 | 0 | 5 | |
| Exon_19 | 178782539 | 178782602 | near Z-disk | 64 | 100 | 100 | 100 | 0.364466 | 6.80855 | 5.98111 | 2.63148 | 4.23679 | 3.09461 | 19 | 0 | 5 | |
| Exon_20 | 178782212 | 178782427 | near Z-disk | 216 | 100 | 100 | 100 | 0.364553 | 8.65834 | 6.46311 | 2.63148 | 4.23679 | 3.02237 | 25 | 0 | 5 | |
| Exon_21 | 178781121 | 178781263 | near Z-disk | 143 | 100 | 100 | 100 | 0.364559 | 9.57498 | 6.49748 | 2.63148 | 4.23679 | 3.02237 | 25 | 0 | 5 | |
| Exon_22 | 178780000 | 178780205 | near Z-disk | 206 | 100 | 100 | 100 | 0.364559 | 9.60875 | 6.5301 | 2.63148 | 4.23679 | 3.02237 | 25 | 0 | 5 | |
| Exon_23 | 178779229 | 178779462 | near Z-disk | 234 | 99 | 100 | 100 | 0.364576 | 9.60875 | 6.53287 | 2.63148 | 4.23679 | 3.02237 | 23 | 0 | 5 | |
| Exon_24 | 178778874 | 178779118 | near Z-disk | 245 | 99 | 100 | 100 | 0.364576 | 9.60875 | 6.80855 | 2.01836 | 4.23679 | 3.00958 | 23 | 0 | 5 | |
| Exon_25 | 178777704 | 178777975 | near Z-disk | 272 | 100 | 100 | 100 | 5.98111 | 9.60875 | 8.08988 | 1.52327 | 4.23679 | 2.8986 | 21 | 0 | 2 | |
| Exon_26 | 178777420 | 178777584 | near Z-disk | 165 | 100 | 100 | 100 | 5.98111 | 9.60875 | 8.32015 | 1.52327 | 3.09461 | 2.782235 | 18 | 0 | 0 | Exons 26-28 are hiPSI but did not have strong support for an association here. |
| Exon_27 | 178777149 | 178777317 | near Z-disk | 169 | 100 | 100 | 100 | 5.72043 | 9.60875 | 7.44028 | 1.42733 | 3.09461 | 2.57149 | 33 | 0 | 0 | |
| Exon_28 | 178775356 | 178777049 | near Z-disk / I-band | 1694 | 100 | 100 | 100 | 3.07526 | 9.60875 | 6.58224 | 1.42733 | 3.32127 | 2.61158 | 36 | 0 | 0 | |
| Exon_29 | 178774921 | 178775202 | I-band | 282 | 100 | 100 | 100 | 0.364311 | 9.19997 | 6.328365 | 1.42733 | 3.95754 | 2.487445 | 24 | 0 | 3 | |
| Exon_30 | 178774207 | 178774473 | I-band | 267 | 100 | 100 | 100 | 0.364311 | 8.74518 | 6.17669 | 1.42733 | 3.95754 | 2.52691 | 21 | 0 | 3 | |
| Exon_31 | 178773838 | 178774110 | I-band | 273 | 100 | 100 | 100 | 0.364311 | 7.44028 | 5.78372 | 1.42733 | 4.45701 | 2.59168 | 19 | 0 | 4 | |
| Exon_32 | 178773462 | 178773725 | I-band | 264 | 100 | 100 | 100 | 0.364311 | 6.61178 | 4.41022 | 2.01221 | 5.01572 | 3.05715 | 23 | 0 | 9 | |
| Exon_33 | 178773109 | 178773369 | I-band | 261 | 100 | 100 | 100 | 0.364311 | 6.49745 | 4.34662 | 2.05269 | 5.01572 | 3.11302 | 21 | 0 | 9 | |
| Exon_34 | 178771211 | 178771471 | I-band | 261 | 100 | 100 | 100 | 0.364311 | 6.41191 | 4.31198 | 2.05269 | 5.01572 | 3.32127 | 19 | 0 | 9 | |
| Exon_35 | 178770412 | 178770675 | I-band | 264 | 100 | 100 | 100 | 0.364311 | 6.41191 | 4.31198 | 2.05269 | 5.01572 | 3.32127 | 19 | 0 | 9 | |
| Exon_36 | 178770060 | 178770320 | I-band | 261 | 100 | 100 | 100 | 0.364311 | 6.24482 | 3.82399 | 2.05269 | 5.01572 | 3.454425 | 18 | 0 | 9 | |
| Exon_37 | 178769679 | 178769939 | I-band | 261 | 100 | 100 | 100 | 0.364311 | 6.62021 | 4.3293 | 2.05269 | 5.08014 | 3.77824 | 20 | 0 | 12 | |
| Exon_38 | 178768673 | 178768933 | I-band | 261 | 100 | 100 | 100 | 0.364311 | 6.62021 | 3.323145 | 2.85258 | 5.08014 | 4.54457 | 16 | 0 | 12 | |
| Exon_39 | 178768014 | 178768155 | I-band | 142 | 100 | 100 | 100 | 0.364311 | 6.62021 | 3.323145 | 2.85258 | 5.08014 | 4.54457 | 16 | 0 | 12 | |
| Exon_40 | 178767759 | 178767924 | I-band | 166 | 100 | 100 | 100 | 0.364311 | 6.62021 | 3.31029 | 3.05715 | 5.08014 | 4.63213 | 15 | 0 | 12 | |
| Exon_41 | 178766381 | 178766612 | I-band | 232 | 100 | 100 | 100 | 0.364311 | 6.62021 | 4.31198 | 3.05715 | 5.08014 | 4.45701 | 17 | 0 | 12 | |
| Exon_42 | 178764527 | 178764811 | I-band | 285 | 100 | 100 | 100 | 0.364323 | 6.62021 | 4.75441 | 3.353 | 5.08014 | 4.65191 | 13 | 0 | 10 | |
| Exon_43 | 178764177 | 178764302 | I-band | 126 | 100 | 100 | 100 | 0.364622 | 6.62021 | 5.36996 | 3.353 | 5.08014 | 4.73437 | 11 | 0 | 8 | |
| Exon_44 | 178758984 | 178759172 | I-band | 189 | 100 | 100 | 100 | 0.364638 | 8.12751 | 5.83762 | 2.38702 | 5.08014 | 4.63213 | 13 | 0 | 7 | |
| Exon_45 | 178757542 | 178757916 | | 375 | 5 | 1 | 5 | 0.36424 | 6.62021 | 5.36996 | 0.865433 | 5.08014 | 3.353 | 13 | 0 | 4 | |
| Exon_46 | 178756222 | 178756797 | | 576 | 14 | 4 | 14 | 0.363983 | 26.0276 | 3.48856 | 0.536388 | 4.4248 | 0.873999 | 45 | 13 | 5 | |
| Exon_47 | 178753124 | 178753180 | I-band | 57 | 100 | 100 | 100 | 0.364135 | 26.0276 | 11.9183 | 0.583627 | 4.4248 | 1.70455 | 27 | 13 | 5 | |
| *Exon_48 | 178745320 | 178752039 | | 6455 | 89 | 82 | 89 | NA | NA | NA | NA | NA | NA | NA | NA | NA | |
| Exon_49 | 178739141 | 178741921 | I-band | 2781 | 95 | 100 | 100 | 0.364135 | 34.5191 | 19.749 | 0.583627 | 5.98104 | 3.76987 | 54 | 36 | 29 | |
| Exon_50 | 178738082 | 178738360 | I-band | 279 | 100 | 100 | 100 | 6.80346 | 34.5191 | 24.9887 | 2.67627 | 5.98104 | 5.605135 | 20 | 16 | 17 | |
| Exon_51 | 178735511 | 178736074 | I-band | 564 | 46 | 33 | 46 | 0.364793 | 34.5191 | 22.3284 | 1.07921 | 5.85339 | 5.50925 | 18 | 13 | 14 | |
| Exon_52 | 178734707 | 178734988 | I-band | 282 | 29 | 15 | 29 | 0.364793 | 25.3483 | 16.2182 | 1.07921 | 5.85339 | 4.44008 | 12 | 7 | 8 | |
| Exon_53 | 178734328 | 178734606 | I-band | 279 | 23 | 10 | 23 | 0.364793 | 20.3213 | 11.6865 | 1.07921 | 5.85339 | 3.26775 | 11 | 4 | 5 | |
| Exon_54 | 178733614 | 178733892 | I-band | 279 | 14 | 7 | 14 | 0.364793 | 11.6865 | 3.584142 | 1.07921 | 3.26775 | 2.092815 | 6 | 0 | 0 | |
| Exon_55 | 178733239 | 178733517 | I-band | 279 | 13 | 6 | 13 | 0.364793 | 11.5623 | 4.600245 | 1.07921 | 3.25388 | 2.63525 | 22 | 0 | 0 | |
| Exon_56 | 178732834 | 178733121 | I-band | 288 | 17 | 6 | 17 | 0.365075 | 10.7829 | 6.20526 | 0.889071 | 3.25192 | 2.37513 | 33 | 0 | 0 | |
| Exon_57 | 178732440 | 178732718 | I-band | 279 | 16 | 7 | 16 | 3.95357 | 10.7829 | 6.28757 | 0.889071 | 3.25192 | 2.05703 | 36 | 0 | 0 | |
| Exon_58 | 178732066 | 178732347 | I-band | 282 | 20 | 7 | 20 | 0.364541 | 10.7829 | 6.003995 | 0.889071 | 2.98644 | 1.927585 | 36 | 0 | 0 | |
| Exon_59 | 178731693 | 178731971 | I-band | 279 | 21 | 8 | 21 | 0.364406 | 10.7829 | 5.5565 | 0.889071 | 2.98644 | 1.751815 | 36 | 0 | 0 | |
| Exon_60 | 178731305 | 178731583 | I-band | 279 | 35 | 20 | 35 | 0.364406 | 10.7829 | 5.378595 | 0.889071 | 2.98644 | 1.7065 | 34 | 0 | 0 | |
| Exon_61 | 178730925 | 178731203 | I-band | 279 | 51 | 35 | 51 | 0.364406 | 10.7829 | 5.45903 | 0.889071 | 2.98644 | 1.67635 | 33 | 0 | 0 | |
| Exon_62 | 178730505 | 178730792 | I-band | 288 | 50 | 32 | 50 | 0.364081 | 10.7829 | 5.20538 | 0.889071 | 2.98644 | 1.76698 | 37 | 0 | 0 | |
| Exon_63 | 178730093 | 178730371 | I-band | 279 | 16 | 6 | 16 | 0.364081 | 10.1687 | 4.74702 | 0.889071 | 2.70013 | 1.6653 | 31 | 0 | 0 | |
| Exon_64 | 178729664 | 178729945 | I-band | 282 | 21 | 7 | 21 | 0.364081 | 10.1687 | 4.38409 | 0.889071 | 2.74338 | 1.6653 | 31 | 0 | 0 | |
| Exon_65 | 178729288 | 178729566 | I-band | 279 | 22 | 8 | 22 | 0.364081 | 10.1687 | 0.364435 | 0.889071 | 3.37246 | 1.7694 | 35 | 0 | 0 | |
| Exon_66 | 178728891 | 178729169 | I-band | 279 | 13 | 4 | 13 | 0.364081 | 9.047 | 0.364245 | 0.889071 | 3.37246 | 1.98114 | 29 | 0 | 0 | |
| Exon_67 | 178728500 | 178728778 | I-band | 279 | 18 | 7 | 18 | 0.364081 | 7.63683 | 0.364199 | 1.20236 | 3.37246 | 2.51013 | 31 | 0 | 0 | |
| Exon_68 | 178728110 | 178728397 | I-band | 288 | 17 | 6 | 17 | 0.364081 | 0.364558 | 0.364197 | 1.15935 | 3.37246 | 2.385315 | 30 | 0 | 0 | |
| Exon_69 | 178727585 | 178727863 | I-band | 279 | 19 | 6 | 19 | 0.364081 | 0.365011 | 0.3642615 | 0.946357 | 3.37246 | 2.20968 | 32 | 0 | 0 | |
| Exon_70 | 178727090 | 178727371 | I-band | 282 | 69 | 52 | 69 | 0.364096 | 0.365011 | 0.364539 | 0.624563 | 3.37246 | 1.30203 | 35 | 0 | 0 | |
| Exon_71 | 178725768 | 178726046 | I-band | 279 | 61 | 46 | 61 | 0.364096 | 6.37245 | 0.364946 | 0.624563 | 3.37246 | 1.16097 | 35 | 0 | 0 | |
| Exon_72 | 178725368 | 178725649 | I-band | 282 | 59 | 46 | 59 | 0.364278 | 6.37245 | 0.364967 | 0.624563 | 3.36358 | 1.076035 | 38 | 0 | 0 | |
| Exon_73 | 178724260 | 178724538 | I-band | 279 | 49 | 26 | 49 | 0.364527 | 6.37245 | 0.3649995 | 0.624563 | 1.33189 | 1.075405 | 38 | 0 | 0 | |
| Exon_74 | 178723856 | 178724143 | I-band | 288 | 51 | 25 | 51 | 0.364558 | 6.37245 | 4.32894 | 0.624563 | 1.42094 | 1.08115 | 40 | 0 | 0 | |
| Exon_75 | 178723418 | 178723696 | I-band | 279 | 55 | 34 | 55 | 0.364961 | 6.37245 | 4.443685 | 0.624563 | 1.42094 | 1.059855 | 34 | 0 | 0 | |

Table S2 - TTN exons and metadata

| Exon | hg38 start | hg38 end | band | Coding length | GTEx PSI | DCM PSI | Max PSI | CM: min OR for overlapping windows | CM: max OR for overlapping windows | CM: median OR for overlapping windows | Afib: min OR for overlapping windows | Afib: max OR for overlapping windows | Afib: median OR for overlapping windows | n windows | CM: n sig windows | Afib: n sig windows | Note |
|---|---|---|---|---|---|---|---|---|---|---|---|---|---|---|---|---|---|
| Exon_76 | 178723046 | 178723324 | I-band | 279 | 55 | 32 | 55 | 0.364961 | 6.37245 | 4.47721 | 0.624563 | 1.92783 | 1.082945 | 34 | 0 | 0 | |
| Exon_77 | 178722659 | 178722937 | I-band | 279 | 76 | 56 | 76 | 0.364961 | 6.37245 | 4.60204 | 0.648299 | 1.92783 | 1.09194 | 31 | 0 | 0 | |
| Exon_78 | 178722259 | 178722546 | I-band | 288 | 61 | 33 | 61 | 3.52737 | 6.37245 | 4.729165 | 0.960692 | 1.92783 | 1.117665 | 32 | 0 | 0 | |
| Exon_79 | 178721847 | 178722134 | I-band | 288 | 65 | 35 | 65 | 3.52737 | 5.60823 | 4.67483 | 0.960692 | 2.15433 | 1.1221 | 29 | 0 | 0 | |
| Exon_80 | 178720921 | 178721202 | I-band | 282 | 76 | 41 | 76 | 3.52737 | 6.65602 | 4.66501 | 0.960692 | 2.57413 | 1.24258 | 26 | 0 | 0 | |
| Exon_81 | 178720385 | 178720663 | I-band | 279 | 75 | 41 | 75 | 3.52737 | 6.65602 | 4.729165 | 1.04942 | 3.39251 | 1.48575 | 28 | 0 | 0 | |
| Exon_82 | 178719983 | 178720264 | I-band | 282 | 76 | 46 | 76 | 3.52737 | 6.65602 | 4.7781 | 1.08052 | 3.39251 | 1.54205 | 29 | 0 | 0 | |
| Exon_83 | 178719554 | 178719832 | I-band | 279 | 77 | 48 | 77 | 3.52737 | 6.65602 | 4.848885 | 1.09194 | 3.39251 | 1.63684 | 24 | 0 | 0 | |
| Exon_84 | 178719164 | 178719451 | I-band | 288 | 37 | 7 | 37 | 3.52737 | 6.65602 | 4.9654 | 1.10336 | 3.39251 | 1.91048 | 25 | 0 | 0 | |
| Exon_85 | 178718695 | 178718973 | I-band | 279 | 29 | 3 | 29 | 3.52737 | 6.65602 | 5.0021 | 1.352 | 3.39251 | 1.92783 | 23 | 0 | 0 | |
| Exon_86 | 178718322 | 178718600 | I-band | 279 | 28 | 4 | 28 | 3.79937 | 6.65602 | 4.9654 | 1.47743 | 3.39251 | 2.35454 | 27 | 0 | 0 | |
| Exon_87 | 178717943 | 178718221 | I-band | 279 | 35 | 4 | 35 | 0.364134 | 6.65602 | 4.81148 | 1.47743 | 3.39251 | 2.46046 | 28 | 0 | 0 | |
| Exon_88 | 178717523 | 178717810 | I-band | 288 | 33 | 4 | 33 | 0.363987 | 6.65602 | 4.63133 | 1.63111 | 3.39251 | 2.52324 | 25 | 0 | 0 | |
| Exon_89 | 178717095 | 178717382 | I-band | 288 | 37 | 8 | 37 | 0.36391 | 6.55443 | 2.81271 | 1.4045 | 3.39251 | 2.390825 | 34 | 0 | 0 | |
| Exon_90 | 178715493 | 178715774 | I-band | 282 | 79 | 59 | 79 | 0.363854 | 5.70467 | 0.36417 | 1.4045 | 3.01667 | 2.30912 | 31 | 0 | 0 | |
| Exon_91 | 178714986 | 178715264 | I-band | 279 | 81 | 54 | 81 | 0.363854 | 5.70467 | 0.364139 | 0.941593 | 3.01667 | 2.03829 | 30 | 0 | 0 | |
| Exon_92 | 178714292 | 178714573 | I-band | 282 | 83 | 60 | 83 | 0.363854 | 2.84316 | 0.3640465 | 0.542318 | 2.78683 | 1.68273 | 24 | 0 | 0 | |
| Exon_93 | 178713897 | 178714175 | I-band | 279 | 85 | 62 | 85 | 0.363854 | 2.78226 | 0.364017 | 0.542318 | 2.70023 | 1.68022 | 23 | 0 | 0 | |
| Exon_94 | 178713085 | 178713372 | I-band | 288 | 85 | 60 | 85 | 0.363854 | 2.78226 | 0.364017 | 0.542318 | 2.70023 | 1.68022 | 23 | 0 | 0 | |
| Exon_95 | 178712697 | 178712975 | I-band | 279 | 85 | 64 | 85 | 0.363854 | 0.364184 | 0.3640145 | 0.542318 | 2.46046 | 1.67848 | 22 | 0 | 0 | |
| Exon_96 | 178712315 | 178712593 | I-band | 279 | 85 | 63 | 85 | 0.363854 | 0.364335 | 0.364076 | 0.440331 | 2.46046 | 1.63801 | 25 | 0 | 0 | |
| Exon_97 | 178711944 | 178712222 | I-band | 279 | 84 | 61 | 84 | 0.363854 | 0.364397 | 0.364084 | 0.440331 | 1.81738 | 1.4326 | 20 | 0 | 0 | |
| Exon_98 | 178711062 | 178711349 | I-band | 288 | 87 | 66 | 87 | 0.363854 | 0.364397 | 0.364103 | 0.440331 | 1.81738 | 1.28338 | 20 | 0 | 0 | |
| Exon_99 | 178710635 | 178710922 | I-band | 288 | 87 | 66 | 87 | 0.363854 | 0.364397 | 0.3640935 | 0.440331 | 2.15556 | 1.167835 | 18 | 0 | 0 | |
| Exon_100 | 178709566 | 178709856 | I-band | 291 | 87 | 69 | 87 | 0.363854 | 0.364397 | 0.364095 | 0.440331 | 2.15556 | 1.02834 | 15 | 0 | 0 | |
| Exon_101 | 178707526 | 178707813 | I-band | 288 | 90 | 72 | 90 | 0.363872 | 5.62571 | 0.364335 | 0.440331 | 2.28262 | 1.28149 | 19 | 0 | 0 | |
| Exon_102 | 178706862 | 178706954 | I-band | 93 | 88 | 72 | 88 | 0.364012 | 5.62571 | 3.80435 | 0.440331 | 2.98583 | 2.270975 | 34 | 0 | 0 | |
| Exon_103 | 178706454 | 178706739 | I-band | 286 | 89 | 75 | 89 | 0.363768 | 6.53521 | 4.06495 | 0.440331 | 2.98583 | 2.26668 | 37 | 0 | 0 | |
| Exon_104 | 178705174 | 178705357 | I-band | 184 | 90 | 76 | 90 | 0.363754 | 6.53521 | 4.0655 | 0.948612 | 2.98583 | 2.3942 | 30 | 0 | 0 | |
| Exon_105 | 178704877 | 178704966 | I-band | 90 | 90 | 75 | 90 | 0.363754 | 6.53521 | 4.017205 | 0.948612 | 2.98583 | 2.62368 | 26 | 0 | 0 | |
| Exon_106 | 178704510 | 178704777 | I-band | 268 | 90 | 70 | 90 | 0.363754 | 6.53521 | 4.06495 | 0.948612 | 2.98583 | 2.686745 | 22 | 0 | 0 | |
| Exon_107 | 178704147 | 178704407 | I-band | 261 | 90 | 73 | 90 | 0.363754 | 6.53521 | 4.10966 | 0.948612 | 2.98583 | 2.70714 | 20 | 0 | 0 | |
| Exon_108 | 178702454 | 178702663 | I-band | 210 | 91 | 74 | 91 | 0.363754 | 6.53521 | 4.06495 | 0.948612 | 2.98583 | 2.70714 | 17 | 0 | 0 | This cluster of isolated hiPSI exons lacks power to test for an association (only 13 TTNtv carriers in exons 108-113). However, the exons from from 91-118 had |
| Exon_109 | 178702168 | 178702245 | I-band | 78 | 92 | 74 | 92 | 0.363754 | 6.53521 | 3.96946 | 0.948612 | 2.98583 | 1.37719 | 13 | 0 | 0 | |
| Exon_110 | 178702040 | 178702066 | I-band | 27 | 91 | 74 | 91 | 0.363754 | 6.53521 | 4.325205 | 0.948612 | 2.96037 | 1.3623 | 12 | 0 | 0 | |
| Exon_111 | 178701528 | 178701587 | I-band | 60 | 93 | 80 | 93 | 0.363754 | 6.53521 | 3.88726 | 0.699207 | 2.96037 | 1.34741 | 13 | 0 | 0 | |
| Exon_112 | 178701120 | 178701203 | I-band | 84 | 93 | 82 | 93 | 0.363754 | 6.53521 | 3.74268 | 0.699207 | 2.92788 | 1.34546 | 13 | 0 | 0 | |
| Exon_113 | 178698843 | 178698914 | I-band | 72 | 91 | 80 | 91 | 0.363754 | 6.53521 | 2.6932045 | 0.699207 | 2.12009 | 1.3394 | 10 | 0 | 0 | |
| Exon_114 | 178697121 | 178697168 | I-band | 48 | 90 | 75 | 90 | 0.363754 | 6.53521 | 0.364145 | 0.699207 | 2.12009 | 1.33733 | 11 | 0 | 0 | |
| Exon_115 | 178695865 | 178696269 | I-band | 405 | 85 | 74 | 85 | 0.363754 | 5.75056 | 0.3644545 | 0.699207 | 1.37719 | 1.172235 | 20 | 0 | 0 | |
| Exon_116 | 178695348 | 178695410 | I-band | 63 | 88 | 75 | 88 | 0.363784 | 0.364531 | 0.36445 | 0.699207 | 1.35239 | 1.12764 | 13 | 0 | 0 | |
| Exon_117 | 178694829 | 178694906 | I-band | 78 | 85 | 63 | 85 | 0.363784 | 0.364791 | 0.36448 | 0.699207 | 1.51198 | 1.172235 | 16 | 0 | 0 | |
| Exon_118 | 178694599 | 178694676 | I-band | 78 | 85 | 59 | 85 | 0.364145 | 0.365091 | 0.364518 | 0.776769 | 1.51198 | 1.16015 | 17 | 0 | 0 | |
| Exon_119 | 178693922 | 178694008 | I-band | 87 | 79 | 57 | 79 | 0.364419 | 0.365091 | 0.3645285 | 0.900383 | 1.51198 | 1.187515 | 14 | 0 | 0 | |
| Exon_120 | 178693609 | 178693689 | I-band | 81 | 79 | 56 | 79 | 0.36445 | 0.365091 | 0.364531 | 0.900383 | 1.51198 | 1.19071 | 13 | 0 | 0 | |
| Exon_121 | 178692497 | 178692580 | I-band | 84 | 79 | 58 | 79 | 0.364459 | 0.365091 | 0.3646475 | 0.900383 | 1.51198 | 1.187515 | 12 | 0 | 0 | |
| Exon_122 | 178692016 | 178692099 | I-band | 84 | 76 | 54 | 76 | 0.364518 | 0.365091 | 0.364768 | 0.900383 | 1.51198 | 1.16015 | 9 | 0 | 0 | |
| Exon_123 | 178689813 | 178689896 | I-band | 84 | 88 | 78 | 88 | 0.364531 | 3.12907 | 0.364925 | 0.900383 | 1.51198 | 1.13072 | 8 | 0 | 0 | |
| Exon_124 | 178689515 | 178689595 | I-band | 81 | 45 | 22 | 45 | 0.364186 | 0.365091 | 0.364253 | 0.900383 | 1.47691 | 1.35811 | 9 | 0 | 0 | |
| Exon_125 | 178689290 | 178689373 | I-band | 84 | 46 | 25 | 46 | 0.363961 | 1.85918 | 0.364121 | 0.900383 | 1.68844 | 1.3498 | 19 | 0 | 0 | |
| Exon_126 | 178689053 | 178689136 | I-band | 84 | 37 | 15 | 37 | 0.363961 | 7.04396 | 0.364121 | 0.727386 | 1.68844 | 1.21749 | 19 | 0 | 0 | |
| Exon_127 | 178688677 | 178688778 | I-band | 102 | 33 | 15 | 33 | 0.363961 | 7.04396 | 0.364121 | 0.706813 | 1.68844 | 1.1612 | 19 | 0 | 0 | |
| Exon_128 | 178688111 | 178688224 | I-band | 114 | 47 | 28 | 47 | 0.363961 | 7.04396 | 1.79318 | 0.622231 | 1.68844 | 1.06877 | 24 | 0 | 0 | |
| Exon_129 | 178685518 | 178685598 | I-band | 81 | 48 | 28 | 48 | 0.364001 | 7.04396 | 3.28417 | 0.614675 | 1.44279 | 0.874102 | 23 | 0 | 0 | |
| Exon_130 | 178685253 | 178685330 | I-band | 78 | 38 | 9 | 38 | 0.364106 | 7.04396 | 3.342695 | 0.604432 | 1.20867 | 0.8497895 | 22 | 0 | 0 | |
| Exon_131 | 178684906 | 178684989 | I-band | 84 | 25 | 10 | 25 | 0.364128 | 7.04396 | 3.28417 | 0.604432 | 1.1612 | 0.763915 | 25 | 0 | 0 | |
| Exon_132 | 178684666 | 178684749 | I-band | 84 | 25 | 6 | 25 | 2.47377 | 4.69978 | 3.40122 | 0.604432 | 0.885303 | 0.706813 | 21 | 0 | 0 | |
| Exon_133 | 178684330 | 178684413 | I-band | 84 | 21 | 10 | 21 | 2.47377 | 6.15751 | 3.28417 | 0.481694 | 0.870072 | 0.622231 | 19 | 0 | 0 | |
| Exon_134 | 178683999 | 178684082 | I-band | 84 | 25 | 12 | 25 | 2.47377 | 6.15751 | 3.262295 | 0.472065 | 0.814544 | 0.616551 | 16 | 0 | 0 | |
| Exon_135 | 178683211 | 178683291 | I-band | 81 | 43 | 26 | 43 | 2.47377 | 6.15751 | 3.279935 | 0.472065 | 0.814544 | 0.616551 | 18 | 0 | 0 | |
| Exon_136 | 178682697 | 178682903 | I-band | 207 | 86 | 76 | 86 | 2.47377 | 6.48666 | 5.6883 | 0.472065 | 1.32905 | 0.804509 | 30 | 0 | 0 | |
| Exon_137 | 178681661 | 178681738 | I-band | 78 | 87 | 77 | 87 | 2.47377 | 6.48666 | 5.83167 | 0.472065 | 1.32905 | 0.849467 | 25 | 0 | 0 | |
| Exon_138 | 178681376 | 178681450 | I-band | 75 | 7 | 4 | 7 | 2.47377 | 6.48666 | 5.830565 | 0.472065 | 1.32905 | 0.933856 | 28 | 0 | 0 | |
| Exon_139 | 178681079 | 178681171 | I-band | 93 | 5 | 1 | 5 | 3.30766 | 6.48666 | 5.830565 | 0.472065 | 1.32905 | 1.0321325 | 28 | 0 | 0 | |
| Exon_140 | 178680254 | 178680331 | I-band | 78 | 5 | 1 | 5 | 3.10359 | 6.48666 | 5.83389 | 0.821454 | 1.32905 | 1.093955 | 24 | 0 | 0 | |
| Exon_141 | 178679894 | 178680055 | I-band | 162 | 5 | 1 | 5 | 3.10359 | 6.48666 | 5.83167 | 0.821454 | 1.32905 | 1.09906 | 23 | 0 | 0 | |
| Exon_142 | 178679599 | 178679682 | I-band | 84 | 6 | 1 | 6 | 3.10359 | 6.48666 | 5.82946 | 0.821454 | 1.32905 | 1.10761 | 21 | 0 | 0 | |
| Exon_143 | 178679339 | 178679416 | I-band | 78 | 6 | 2 | 6 | 3.10359 | 6.48666 | 5.82946 | 0.821454 | 1.32905 | 1.10761 | 21 | 0 | 0 | |
| Exon_144 | 178678747 | 178678830 | I-band | 84 | 88 | 74 | 88 | 3.10359 | 6.48666 | 5.78734 | 0.821454 | 1.32905 | 1.10985 | 20 | 0 | 0 | |
| Exon_145 | 178678414 | 178678497 | I-band | 84 | 83 | 69 | 83 | 3.10359 | 6.25359 | 5.74522 | 0.821454 | 1.28223 | 1.09906 | 19 | 0 | 0 | |
| Exon_146 | 178678125 | 178678208 | I-band | 84 | 86 | 68 | 86 | 3.10359 | 6.23508 | 5.73958 | 0.821454 | 1.22253 | 1.08885 | 17 | 0 | 0 | |
| Exon_147 | 178677621 | 178677917 | I-band | 297 | 10 | 2 | 10 | 0.364277 | 5.93715 | 5.69246 | 0.821454 | 1.53033 | 1.07047 | 15 | 0 | 0 | |
| Exon_148 | 178677201 | 178677287 | | 87 | 7 | 2 | 7 | 3.10359 | 5.93715 | 5.603295 | 0.821454 | 1.20307 | 0.934695 | 12 | 0 | 0 | |
| Exon_149 | 178675921 | 178675995 | I-band | 75 | 11 | 3 | 11 | 0.362662 | 5.93715 | 5.28484 | 0.821454 | 1.46338 | 0.875595 | 11 | 0 | 0 | |
| Exon_150 | 178675671 | 178675754 | | 84 | 5 | 1 | 5 | 0.362662 | 5.92293 | 3.85927 | 0.836973 | 1.46338 | 0.993795 | 9 | 0 | 0 | |

Table S2 - TTN exons and metadata

| Exon | hg38 start | hg38 end | band | Coding length | GTEx PSI | DCM PSI | Max PSI | CM: min OR for overlapping windows | CM: max OR for overlapping windows | CM: median OR for overlapping windows | Afib: min OR for overlapping windows | Afib: max OR for overlapping windows | Afib: median OR for overlapping windows | n windows | CM: n sig windows | Afib: n sig windows | Note |
|---|---|---|---|---|---|---|---|---|---|---|---|---|---|---|---|---|---|
| Exon_151 | 178675039 | 178675113 | I-band | 75 | 21 | 20 | 21 | 0.362662 | 5.51413 | 3.693855 | 0.854527 | 1.46338 | 1.0321325 | 8 | 0 | 0 | |
| Exon_152 | 178674314 | 178674409 | I-band | 96 | 79 | 54 | 79 | 0.362662 | 4.06172 | 3.30766 | 0.854527 | 1.46338 | 0.875595 | 5 | 0 | 0 | |
| Exon_153 | 178673633 | 178673710 | I-band | 78 | 75 | 48 | 75 | 0.362662 | 7.85501 | 3.79508 | 0.854527 | 1.85333 | 1.430235 | 8 | 0 | 0 | |
| Exon_154 | 178672635 | 178672703 | I-band | 69 | 27 | 10 | 27 | 0.362662 | 8.8683 | 5.6105 | 1.39709 | 2.63359 | 1.981645 | 18 | 0 | 0 | |
| Exon_155 | 178672407 | 178672481 | I-band | 75 | 14 | 3 | 14 | 3.77359 | 9.3069 | 5.71818 | 1.3159 | 2.63359 | 1.97172 | 19 | 0 | 0 | |
| Exon_156 | 178671971 | 178672267 | I-band | 297 | 11 | 2 | 11 | 3.77359 | 9.3069 | 5.758675 | 1.3159 | 2.63359 | 1.981645 | 18 | 0 | 0 | |
| Exon_157 | 178671090 | 178671170 | I-band | 81 | 16 | 5 | 16 | 3.77359 | 9.3069 | 5.478375 | 1.3159 | 2.63359 | 2.148045 | 14 | 0 | 0 | |
| Exon_158 | 178670218 | 178670295 | I-band | 78 | 66 | 30 | 66 | 3.77359 | 9.3069 | 5.204335 | 1.3159 | 2.63359 | 2.23411 | 12 | 0 | 0 | |
| Exon_159 | 178669592 | 178669675 | | 84 | 6 | 1 | 6 | 3.77359 | 9.3069 | 4.95474 | 1.3159 | 2.55881 | 2.23289 | 11 | 0 | 0 | |
| Exon_160 | 178669373 | 178669447 | | 75 | 7 | 1 | 7 | 0.365016 | 9.3069 | 4.93304 | 1.3159 | 2.55881 | 2.208525 | 12 | 0 | 0 | |
| Exon_161 | 178667638 | 178667721 | | 84 | 6 | 3 | 6 | 0.365016 | 9.3069 | 4.216175 | 1.3159 | 2.55881 | 2.07794 | 8 | 0 | 0 | |
| Exon_162 | 178667442 | 178667525 | | 84 | 5 | 2 | 5 | 0.365016 | 9.3069 | 3.88098 | 1.3159 | 2.55881 | 2.28968 | 6 | 0 | 0 | |
| Exon_163 | 178667236 | 178667319 | | 84 | 5 | 1 | 5 | 0.364667 | 4.95474 | 0.364851 | 0.786698 | 2.3952 | 1.086025 | 14 | 0 | 0 | |
| Exon_164 | 178666824 | 178666901 | | 78 | 5 | 0 | 5 | 0.364358 | 0.364967 | 0.364705 | 0.418035 | 1.16272 | 0.802965 | 17 | 0 | 0 | |
| Exon_165 | 178665708 | 178665791 | | 84 | 6 | 3 | 6 | 0.364358 | 0.364882 | 0.3646975 | 0.418035 | 1.16272 | 0.796669 | 16 | 0 | 0 | |
| Exon_166 | 178665377 | 178665460 | | 84 | 5 | 3 | 5 | 0.364358 | 0.364857 | 0.3646785 | 0.418035 | 1.16272 | 0.7885355 | 14 | 0 | 0 | |
| Exon_167 | 178664852 | 178664926 | | 75 | 5 | 2 | 5 | 0.364358 | 0.364857 | 0.364667 | 0.418035 | 1.09757 | 0.786698 | 13 | 0 | 0 | |
| Exon_168 | 178664654 | 178664737 | | 84 | 9 | 3 | 9 | 0.364358 | 0.364857 | 0.364423 | 0.418035 | 1.03572 | 0.448899 | 11 | 0 | 0 | |
| Exon_169 | 178664460 | 178664537 | | 78 | 7 | 2 | 7 | 0.363769 | 0.36486 | 0.36469 | 0.335349 | 1.23564 | 0.420193 | 15 | 0 | 0 | |
| Exon_170 | 178664015 | 178664098 | | 84 | 8 | 2 | 8 | 0.364393 | 0.365061 | 0.364837 | 0.335349 | 0.420503 | 0.340161 | 14 | 0 | 0 | |
| Exon_171 | 178663819 | 178663902 | | 84 | 7 | 1 | 7 | 0.364423 | 7.84001 | 0.364953 | 0.335349 | 1.03642 | 0.340527 | 17 | 0 | 0 | |
| Exon_172 | 178663627 | 178663710 | | 84 | 5 | 1 | 5 | 0.364727 | 8.18366 | 0.3649785 | 0.335587 | 1.11074 | 0.3409215 | 12 | 0 | 0 | |
| Exon_173 | 178663433 | 178663516 | | 84 | 28 | 7 | 28 | 0.364727 | 8.40763 | 0.3650275 | 0.340527 | 1.15837 | 0.341944 | 10 | 0 | 0 | |
| Exon_174 | 178663266 | 178663349 | | 84 | 10 | 3 | 10 | 0.364727 | 8.42729 | 2.5839105 | 0.340905 | 1.15837 | 0.5435365 | 10 | 0 | 0 | |
| Exon_175 | 178662966 | 178663055 | I-band | 90 | 51 | 51 | 51 | 0.364727 | 8.42729 | 6.321385 | 0.340905 | 1.15837 | 0.890666 | 8 | 0 | 0 | |
| Exon_176 | 178662729 | 178662812 | | 84 | 4 | 1 | 4 | 0.364727 | 8.42729 | 6.321385 | 0.340905 | 1.15837 | 0.890666 | 8 | 0 | 0 | |
| Exon_177 | 178662533 | 178662616 | | 84 | 5 | 6 | 6 | 0.364727 | 8.42729 | 7.83874 | 0.340905 | 2.41697 | 1.07358 | 8 | 0 | 0 | |
| Exon_178 | 178662338 | 178662418 | | 81 | 3 | 2 | 3 | 0.363779 | 8.42729 | 7.83874 | 0.744912 | 2.44187 | 1.14833 | 8 | 0 | 0 | |
| Exon_179 | 178662142 | 178662222 | | 81 | 4 | 1 | 4 | 0.363779 | 8.42729 | 4.1006625 | 1.13829 | 2.54561 | 2.42038 | 6 | 0 | 0 | |
| Exon_180 | 178661948 | 178662028 | | 81 | 4 | 5 | 5 | 0.36352 | 7.83747 | 0.363855 | 0.93904 | 2.54561 | 1.20028 | 15 | 0 | 0 | |
| Exon_181 | 178661759 | 178661842 | | 84 | 1 | 2 | 2 | 0.36352 | 0.364663 | 0.363867 | 0.93904 | 2.54561 | 1.03731 | 12 | 0 | 0 | |
| Exon_182 | 178659172 | 178659255 | I-band | 84 | 11 | 8 | 11 | 0.36352 | 0.364663 | 0.363867 | 0.93904 | 2.54561 | 1.03731 | 12 | 0 | 0 | |
| Exon_183 | 178659005 | 178659088 | I-band | 84 | 8 | 4 | 8 | 0.36352 | 0.364663 | 0.363867 | 0.93904 | 2.54561 | 1.03731 | 12 | 0 | 0 | |
| Exon_184 | 178658705 | 178658794 | | 90 | 35 | 22 | 35 | 0.36352 | 0.364663 | 0.363913 | 0.93904 | 2.54561 | 1.02707 | 13 | 0 | 0 | |
| Exon_185 | 178658468 | 178658551 | | 84 | 3 | 1 | 3 | 0.36352 | 0.364663 | 0.364297 | 0.855665 | 2.07054 | 0.967299 | 17 | 0 | 0 | |
| Exon_186 | 178658272 | 178658355 | | 84 | 4 | 7 | 7 | 0.36352 | 0.364663 | 0.364297 | 0.855665 | 1.26708 | 0.962468 | 17 | 0 | 0 | |
| Exon_187 | 178658077 | 178658157 | | 81 | 3 | 5 | 5 | 0.363727 | 6.5729 | 0.364583 | 0.637046 | 1.26708 | 0.93904 | 19 | 0 | 0 | |
| Exon_188 | 178657881 | 178657961 | | 81 | 4 | 3 | 4 | 0.363821 | 6.67805 | 0.364636 | 0.637046 | 1.26708 | 0.940809 | 20 | 0 | 0 | |
| Exon_189 | 178657687 | 178657767 | | 81 | 4 | 7 | 7 | 0.363913 | 6.67805 | 4.28597 | 0.637046 | 1.38659 | 0.942578 | 29 | 0 | 0 | |
| Exon_190 | 178657498 | 178657581 | | 84 | 1 | 3 | 3 | 0.364225 | 6.67805 | 4.49657 | 0.635975 | 1.38659 | 0.894436 | 31 | 0 | 0 | |
| Exon_191 | 178654912 | 178654995 | | 84 | 12 | 4 | 12 | 0.364225 | 6.67805 | 4.60429 | 0.635975 | 1.77839 | 0.884205 | 29 | 0 | 0 | |
| Exon_192 | 178654745 | 178654828 | | 84 | 10 | 2 | 10 | 0.364225 | 6.67805 | 4.60429 | 0.635975 | 1.38659 | 0.875797 | 29 | 0 | 0 | |
| Exon_193 | 178654445 | 178654534 | | 90 | 42 | 13 | 42 | 0.364225 | 6.67805 | 4.683335 | 0.635975 | 1.38659 | 0.865731 | 28 | 0 | 0 | |
| Exon_194 | 178654208 | 178654291 | | 84 | 5 | 0 | 5 | 0.364225 | 6.67805 | 4.78496 | 0.635975 | 1.38659 | 0.8074915 | 26 | 0 | 0 | |
| Exon_195 | 178654012 | 178654095 | | 84 | 8 | 5 | 8 | 0.364225 | 6.67805 | 4.55043 | 0.589375 | 1.38659 | 0.686791 | 30 | 0 | 0 | |
| Exon_196 | 178653817 | 178653897 | | 81 | 10 | 3 | 10 | 2.88138 | 6.67805 | 4.55043 | 0.589375 | 1.38659 | 0.674475 | 26 | 0 | 0 | |
| Exon_197 | 178653621 | 178653701 | | 81 | 8 | 2 | 8 | 0.364464 | 6.67805 | 4.47902 | 0.589375 | 1.38659 | 0.674475 | 26 | 0 | 0 | |
| Exon_198 | 178653427 | 178653507 | | 81 | 7 | 7 | 7 | 0.364464 | 6.67805 | 4.43508 | 0.589375 | 1.38659 | 0.686791 | 26 | 0 | 0 | |
| Exon_199 | 178653238 | 178653321 | | 84 | 5 | 3 | 5 | 0.364457 | 6.67805 | 4.35486 | 0.589375 | 1.38659 | 0.674475 | 28 | 0 | 0 | |
| Exon_200 | 178653041 | 178653124 | | 84 | 6 | 2 | 6 | 0.364433 | 6.02922 | 4.11929 | 0.589375 | 1.2887 | 0.648256 | 25 | 0 | 0 | |
| Exon_201 | 178652848 | 178652931 | | 84 | 3 | 0 | 3 | 0.364333 | 6.02922 | 3.439685 | 0.589375 | 1.21507 | 0.630232 | 28 | 0 | 0 | |
| Exon_202 | 178652653 | 178652736 | | 84 | 3 | 0 | 3 | 0.364333 | 4.49657 | 0.364491 | 0.589375 | 0.812005 | 0.622929 | 21 | 0 | 0 | |
| Exon_203 | 178652458 | 178652541 | I-band | 84 | 8 | 3 | 8 | 0.364333 | 4.46147 | 0.364483 | 0.589375 | 0.812005 | 0.622338 | 20 | 0 | 0 | |
| Exon_204 | 178652264 | 178652347 | I-band | 84 | 25 | 8 | 25 | 0.364333 | 4.46147 | 0.3644695 | 0.589375 | 0.812005 | 0.622338 | 20 | 0 | 0 | |
| Exon_205 | 178652096 | 178652179 | I-band | 84 | 43 | 6 | 43 | 0.364304 | 4.06339 | 0.364457 | 0.589375 | 0.812005 | 0.620878 | 21 | 0 | 0 | |
| Exon_206 | 178651884 | 178651967 | I-band | 84 | 49 | 5 | 49 | 0.364304 | 3.94775 | 0.3644465 | 0.589375 | 0.812005 | 0.619884 | 20 | 0 | 0 | |
| Exon_207 | 178651666 | 178651749 | I-band | 84 | 39 | 8 | 39 | 0.364304 | 3.46269 | 0.364433 | 0.589375 | 0.812005 | 0.61889 | 19 | 0 | 0 | |
| Exon_208 | 178651453 | 178651536 | I-band | 84 | 32 | 9 | 32 | 0.364304 | 3.46269 | 0.364432 | 0.589375 | 0.812005 | 0.6181185 | 18 | 0 | 0 | |
| Exon_209 | 178651243 | 178651320 | I-band | 78 | 36 | 10 | 36 | 0.364304 | 3.46269 | 0.364431 | 0.589375 | 0.812005 | 0.617347 | 17 | 0 | 0 | |
| Exon_210 | 178650751 | 178650834 | I-band | 84 | 34 | 7 | 34 | 0.364304 | 3.46269 | 0.364431 | 0.589375 | 0.812005 | 0.61889 | 19 | 0 | 0 | |
| Exon_211 | 178650164 | 178650271 | I-band | 108 | 25 | 7 | 25 | 0.364304 | 0.364491 | 0.36442 | 0.601499 | 0.812005 | 0.62085 | 17 | 0 | 0 | |
| Exon_212 | 178649817 | 178649894 | I-band | 78 | 24 | 8 | 24 | 0.363836 | 0.364491 | 0.3643365 | 0.601499 | 1.40672 | 0.653203 | 26 | 0 | 0 | |
| Exon_213 | 178649554 | 178649631 | | 78 | 16 | 3 | 16 | 0.363836 | 0.36445 | 0.364214 | 0.607913 | 1.47526 | 0.829695 | 22 | 0 | 0 | |
| Exon_214 | 178649248 | 178649331 | | 84 | 18 | 2 | 18 | 0.363836 | 12.5086 | 0.3643235 | 0.607913 | 2.61904 | 1.25748 | 28 | 2 | 0 | |
| Exon_215 | 178647381 | 178647464 | | 84 | 18 | 2 | 18 | 0.363836 | 12.5086 | 0.364304 | 0.612864 | 3.28229 | 1.34634 | 25 | 2 | 0 | |
| Exon_216 | 178647064 | 178647144 | | 81 | 10 | 1 | 10 | 0.363836 | 12.5086 | 8.670955 | 0.612864 | 3.37745 | 2.537235 | 36 | 5 | 0 | |
| Exon_217 | 178646485 | 178646559 | | 75 | 14 | 1 | 14 | 0.363836 | 13.3935 | 10.39995 | 0.671142 | 4.55209 | 2.747795 | 44 | 10 | 11 | |
| Exon_218 | 178645920 | 178646030 | I-band | 111 | 23 | 3 | 23 | 0.363836 | 13.3935 | 10.5266 | 0.988248 | 4.55209 | 2.86207 | 42 | 10 | 11 | |
| Exon_219 | 178644548 | 178644616 | I-band | 69 | 17 | 4 | 17 | 0.363862 | 13.3935 | 10.6321 | 0.988248 | 4.55209 | 2.9096 | 41 | 10 | 11 | |
| Exon_220 | 178642237 | 178642317 | I-band | 81 | 96 | 100 | 100 | 0.36389 | 13.3935 | 10.6321 | 1.06017 | 4.55209 | 2.9096 | 41 | 10 | 11 | |
| Exon_221 | 178641241 | 178641315 | I-band | 75 | 95 | 100 | 100 | 0.36389 | 13.3935 | 10.73695 | 1.25541 | 4.55209 | 2.97302 | 40 | 10 | 11 | |
| Exon_222 | 178640541 | 178640630 | I-band | 90 | 95 | 100 | 100 | 0.363956 | 16.2462 | 11.05255 | 1.25541 | 4.85291 | 3.06671 | 42 | 13 | 14 | |
| Exon_223 | 178640048 | 178640110 | I-band | 63 | 95 | 100 | 100 | 0.364028 | 16.2462 | 11.4835 | 1.25541 | 4.85291 | 3.26061 | 38 | 13 | 14 | |
| Exon_224 | 178639699 | 178639788 | I-band | 90 | 96 | 100 | 100 | 0.364028 | 16.8575 | 11.6241 | 1.25541 | 5.04854 | 3.28229 | 39 | 14 | 15 | |
| Exon_225 | 178637369 | 178637419 | I-band | 51 | 73 | 79 | 79 | 0.364051 | 16.8575 | 11.6723 | 1.25541 | 5.04854 | 3.306335 | 38 | 14 | 15 | |

Table S2 - TTN exons and metadata

| Exon | hg38 start | hg38 end | band | Coding length | GTEx PSI | DCM PSI | Max PSI | CM: min OR for overlapping windows | CM: max OR for overlapping windows | CM: median OR for overlapping windows | Afib: min OR for overlapping windows | Afib: max OR for overlapping windows | Afib: median OR for overlapping windows | n windows | CM: n sig windows | Afib: n sig windows | Note |
|---|---|---|---|---|---|---|---|---|---|---|---|---|---|---|---|---|---|
| Exon_226 | 178636398 | 178636799 | I-band | 402 | 99 | 100 | 100 | 0.364051 | 16.8575 | 11.7205 | 1.25541 | 5.04854 | 3.33038 | 39 | 15 | 16 | |
| Exon_227 | 178635963 | 178636241 | I-band | 279 | 99 | 100 | 100 | 0.364054 | 18.0158 | 11.9337 | 1.47526 | 5.04854 | 3.517835 | 44 | 21 | 22 | |
| Exon_228 | 178635440 | 178635715 | I-band | 276 | 98 | 100 | 100 | 8.48696 | 18.0158 | 12.1817 | 2.47712 | 5.04854 | 3.65822 | 43 | 22 | 22 | |
| Exon_229 | 178635165 | 178635304 | I-band | 140 | 98 | 100 | 100 | 8.48696 | 18.0158 | 12.37795 | 2.47712 | 5.04854 | 3.81505 | 40 | 21 | 22 | |
| Exon_230 | 178634723 | 178634849 | I-band | 127 | 97 | 100 | 100 | 8.48696 | 18.0158 | 12.4099 | 2.47712 | 5.04854 | 3.93647 | 39 | 21 | 22 | |
| Exon_231 | 178634366 | 178634629 | I-band | 264 | 98 | 100 | 100 | 8.48696 | 18.0158 | 12.4099 | 2.57253 | 5.04854 | 3.967285 | 38 | 21 | 22 | |
| Exon_232 | 178633817 | 178634083 | I-band | 267 | 98 | 100 | 100 | 9.25593 | 18.0158 | 12.4579 | 2.57253 | 5.04854 | 3.9986 | 35 | 18 | 23 | |
| Exon_233 | 178633413 | 178633676 | I-band | 264 | 98 | 100 | 100 | 9.25593 | 18.0158 | 12.4579 | 2.57253 | 5.04854 | 3.9986 | 35 | 18 | 27 | |
| Exon_234 | 178633187 | 178633326 | I-band | 140 | 97 | 100 | 100 | 9.25593 | 18.0158 | 12.83145 | 2.83374 | 5.04854 | 4.12655 | 32 | 18 | 28 | |
| Exon_235 | 178632918 | 178633044 | I-band | 127 | 97 | 100 | 100 | 9.25593 | 18.0158 | 13.10565 | 2.83374 | 5.04854 | 4.155065 | 30 | 18 | 26 | |
| Exon_236 | 178632526 | 178632792 | I-band | 267 | 98 | 100 | 100 | 9.25593 | 18.0158 | 13.023 | 2.83374 | 5.04854 | 4.15267 | 31 | 18 | 26 | |
| Exon_237 | 178632147 | 178632413 | I-band | 267 | 98 | 100 | 100 | 9.25593 | 18.0158 | 11.7916 | 2.83374 | 5.04854 | 3.69363 | 29 | 14 | 22 | |
| Exon_238 | 178631034 | 178631300 | I-band | 267 | 96 | 100 | 100 | 9.25593 | 18.0158 | 13.0408 | 2.83374 | 5.04854 | 3.665995 | 26 | 13 | 18 | |
| Exon_239 | 178630804 | 178630943 | I-band | 140 | 96 | 100 | 100 | 9.25593 | 18.0158 | 11.1774 | 2.83374 | 5.04854 | 3.6017 | 28 | 13 | 17 | |
| Exon_240 | 178630241 | 178630367 | I-band | 127 | 96 | 100 | 100 | 9.25593 | 18.0158 | 10.82265 | 2.83374 | 5.04854 | 3.566555 | 26 | 11 | 15 | |
| Exon_241 | 178629301 | 178629443 | I-band | 143 | 96 | 100 | 100 | 9.25593 | 18.0158 | 10.79675 | 2.83374 | 4.78446 | 3.55732 | 24 | 9 | 13 | |
| Exon_242 | 178625273 | 178625396 | I-band | 124 | 96 | 100 | 100 | 9.25593 | 17.6111 | 10.6516 | 2.83374 | 4.75546 | 3.43604 | 20 | 5 | 9 | |
| Exon_243 | 178624465 | 178624731 | I-band | 267 | 69 | 59 | 69 | 9.25593 | 17.6111 | 10.6539 | 2.83374 | 4.77127 | 3.48157 | 21 | 6 | 10 | |
| Exon_244 | 178622670 | 178622767 | I-band | 98 | 96 | 100 | 100 | 9.25593 | 19.1772 | 10.82265 | 2.83374 | 5.90561 | 3.65069 | 26 | 11 | 17 | |
| Exon_245 | 178621840 | 178622008 | I-band | 169 | 96 | 100 | 100 | 9.44175 | 19.1772 | 10.8452 | 2.87333 | 5.90561 | 3.67377 | 23 | 11 | 15 | |
| Exon_246 | 178621475 | 178621741 | I-band | 267 | 97 | 100 | 100 | 9.44175 | 28.7265 | 17.3499 | 2.87333 | 6.04248 | 4.839645 | 26 | 16 | 18 | |
| Exon_247 | 178621102 | 178621368 | I-band | 267 | 96 | 100 | 100 | 9.97582 | 37.0287 | 19.0325 | 2.87333 | 6.88245 | 5.216405 | 30 | 25 | 25 | |
| Exon_248 | 178620715 | 178620993 | I-band | 279 | 97 | 100 | 100 | 16.9218 | 37.0287 | 23.9628 | 4.81378 | 7.60699 | 5.61632 | 27 | 27 | 27 | |
| Exon_249 | 178620217 | 178620625 | I-band | 409 | 96 | 100 | 100 | 12.7781 | 37.0287 | 22.47015 | 5.01168 | 7.60699 | 6.04094 | 42 | 42 | 42 | |
| Exon_250 | 178619988 | 178620112 | I-band | 125 | 97 | 100 | 100 | 12.7781 | 37.0287 | 22.5776 | 5.01168 | 7.60699 | 6.31986 | 35 | 35 | 35 | |
| Exon_251 | 178619621 | 178619887 | I-band | 267 | 97 | 100 | 100 | 12.6774 | 37.0287 | 22.5537 | 5.01168 | 7.60699 | 6.362985 | 36 | 36 | 36 | |
| Exon_252 | 178618584 | 178618853 | I-band | 270 | 89 | 100 | 100 | 12.6774 | 37.0287 | 22.4105 | 5.01168 | 7.60699 | 6.41303 | 33 | 33 | 33 | |
| Exon_253 | 178618189 | 178618491 | A-band | 303 | 91 | 100 | 100 | 12.6774 | 37.0287 | 22.3388 | 5.01168 | 7.60699 | 6.43398 | 32 | 32 | 32 | |
| Exon_254 | 178617779 | 178618081 | A-band | 303 | 92 | 100 | 100 | 12.6774 | 37.0287 | 22.2671 | 5.01168 | 7.60699 | 6.40611 | 31 | 31 | 31 | |
| Exon_255 | 178617325 | 178617512 | A-band | 188 | 86 | 100 | 100 | 12.6774 | 24.2113 | 20.10345 | 5.30954 | 7.60699 | 6.46236 | 26 | 26 | 26 | |
| Exon_256 | 178617120 | 178617234 | A-band | 115 | 84 | 100 | 100 | 12.6774 | 24.2113 | 19.857 | 5.32617 | 7.60699 | 6.51861 | 25 | 25 | 25 | |
| Exon_257 | 178616729 | 178617013 | A-band | 285 | 86 | 100 | 100 | 12.6774 | 24.2113 | 19.857 | 5.32617 | 7.60699 | 6.51861 | 25 | 25 | 25 | |
| Exon_258 | 178616479 | 178616630 | A-band | 152 | 86 | 100 | 100 | 0.364422 | 23.9628 | 18.9711 | 3.65215 | 7.60699 | 6.40611 | 25 | 23 | 25 | |
| Exon_259 | 178615641 | 178615788 | A-band | 148 | 85 | 100 | 100 | 0.364408 | 22.882 | 16.7666 | 3.20362 | 7.60699 | 6.215825 | 24 | 20 | 22 | |
| Exon_260 | 178615307 | 178615484 | A-band | 178 | 96 | 100 | 100 | 0.364408 | 21.7939 | 16.0727 | 3.20362 | 7.10816 | 6.01944 | 19 | 15 | 17 | |
| Exon_261 | 178614847 | 178614968 | A-band | 122 | 82 | 100 | 100 | 0.364408 | 19.1116 | 15.61665 | 3.20362 | 7.10816 | 6.02942 | 16 | 12 | 14 | |
| Exon_262 | 178614466 | 178614753 | A-band | 288 | 93 | 100 | 100 | 0.364408 | 16.8946 | 15.3222 | 3.20362 | 7.10816 | 6.01944 | 13 | 9 | 11 | |
| Exon_263 | 178614052 | 178614348 | A-band | 297 | 96 | 100 | 100 | 0.364408 | 15.3909 | 6.520911 | 3.20362 | 7.10816 | 4.835795 | 8 | 4 | 6 | |
| Exon_264 | 178613751 | 178613937 | A-band | 187 | 95 | 100 | 100 | 0.364408 | 13.0874 | 10.1082 | 3.20362 | 4.7874 | 4.20017 | 15 | 5 | 13 | |
| Exon_265 | 178613161 | 178613276 | A-band | 116 | 96 | 100 | 100 | 0.364421 | 20.4762 | 17.6727 | 3.20362 | 7.40387 | 6.57024 | 27 | 20 | 26 | |
| Exon_266 | 178612773 | 178613072 | A-band | 300 | 95 | 100 | 100 | 8.16918 | 22.119 | 18.08675 | 4.02665 | 8.29072 | 6.604895 | 26 | 21 | 26 | |
| Exon_267 | 178612277 | 178612576 | A-band | 300 | 93 | 100 | 100 | 9.63847 | 22.4752 | 18.891 | 4.02665 | 8.65564 | 7.02522 | 37 | 34 | 37 | |
| Exon_268 | 178612057 | 178612162 | A-band | 106 | 93 | 100 | 100 | 12.2797 | 26.1448 | 19.4804 | 4.7874 | 8.65564 | 7.36908 | 35 | 35 | 35 | |
| Exon_269 | 178611758 | 178611954 | A-band | 197 | 94 | 100 | 100 | 17.4781 | 26.8905 | 19.65565 | 6.52685 | 8.65564 | 7.33154 | 36 | 36 | 36 | |
| Exon_270 | 178611372 | 178611677 | A-band | 306 | 94 | 100 | 100 | 17.778 | 26.8905 | 20.1558 | 6.40345 | 8.65564 | 7.36908 | 35 | 35 | 35 | |
| Exon_271 | 178610993 | 178611271 | A-band | 279 | 94 | 100 | 100 | 17.778 | 28.5575 | 21.83535 | 4.5163 | 8.65564 | 7.196515 | 42 | 42 | 42 | |
| Exon_272 | 178610090 | 178610389 | A-band | 300 | 95 | 100 | 100 | 17.778 | 28.5575 | 22.14385 | 4.5163 | 8.65564 | 7.098575 | 42 | 42 | 42 | |
| Exon_273 | 178609684 | 178609986 | A-band | 303 | 95 | 100 | 100 | 17.778 | 28.5575 | 23.5208 | 4.5163 | 8.65564 | 6.52073 | 48 | 48 | 48 | |
| Exon_274 | 178609208 | 178609570 | A-band | 363 | 95 | 100 | 100 | 18.6278 | 28.5575 | 24.1059 | 4.5163 | 8.65564 | 6.34572 | 43 | 43 | 43 | |
| Exon_275 | 178608606 | 178608908 | A-band | 303 | 94 | 100 | 100 | 17.676 | 28.5575 | 24.4141 | 4.5163 | 8.28599 | 5.7088 | 40 | 40 | 40 | |
| Exon_276 | 178608178 | 178608477 | A-band | 300 | 96 | 100 | 100 | 17.5061 | 28.5575 | 25.1699 | 4.50916 | 8.05542 | 5.11768 | 35 | 35 | 35 | |
| Exon_277 | 178607785 | 178608081 | A-band | 297 | 93 | 100 | 100 | 14.8955 | 28.5575 | 24.48915 | 3.18153 | 8.05542 | 4.921105 | 40 | 40 | 38 | |
| Exon_278 | 178607401 | 178607685 | A-band | 285 | 94 | 100 | 100 | 14.8955 | 28.5575 | 24.48915 | 3.18153 | 6.43436 | 4.794115 | 34 | 34 | 32 | |
| Exon_279 | 178607021 | 178607314 | A-band | 294 | 93 | 100 | 100 | 14.8955 | 28.5575 | 24.5436 | 3.18153 | 6.34572 | 4.762 | 35 | 35 | 33 | |
| Exon_280 | 178605414 | 178605713 | A-band | 300 | 93 | 100 | 100 | 14.8955 | 28.5575 | 24.5436 | 3.18153 | 5.82023 | 4.67345 | 33 | 33 | 31 | |
| Exon_281 | 178604987 | 178605295 | A-band | 309 | 93 | 100 | 100 | 14.8955 | 28.5575 | 24.4601 | 3.18153 | 5.82023 | 4.62765 | 35 | 35 | 33 | |
| Exon_282 | 178604708 | 178604898 | A-band | 191 | 93 | 100 | 100 | 14.8955 | 26.9854 | 21.24385 | 3.18153 | 5.82023 | 4.51896 | 30 | 30 | 26 | |
| Exon_283 | 178603876 | 178604305 | A-band | 430 | 94 | 100 | 100 | 14.8955 | 26.3958 | 21.3971 | 3.09284 | 5.82023 | 4.38209 | 31 | 31 | 24 | |
| Exon_284 | 178602282 | 178602590 | A-band | 309 | 89 | 100 | 100 | 14.8955 | 26.0192 | 20.61035 | 3.09284 | 4.97802 | 4.07199 | 26 | 26 | 19 | |
| Exon_285 | 178602002 | 178602150 | A-band | 149 | 90 | 100 | 100 | 14.8955 | 25.8853 | 20.2708 | 3.09284 | 4.8378 | 3.97318 | 25 | 25 | 18 | |
| Exon_286 | 178601882 | 178601914 | A-band | 33 | 87 | 100 | 100 | 14.8955 | 25.8853 | 20.2708 | 3.09284 | 4.8378 | 3.97318 | 25 | 25 | 18 | |
| Exon_287 | 178601658 | 178601787 | A-band | 130 | 90 | 100 | 100 | 14.8955 | 32.9972 | 21.1582 | 2.88716 | 4.72423 | 3.71089 | 39 | 39 | 26 | |
| Exon_288 | 178601265 | 178601564 | A-band | 300 | 91 | 100 | 100 | 14.8955 | 32.9972 | 21.3971 | 2.88716 | 4.72423 | 3.67768 | 35 | 35 | 22 | |
| Exon_289 | 178600854 | 178601171 | A-band | 318 | 89 | 100 | 100 | 14.8955 | 32.9972 | 21.57035 | 2.88716 | 4.72423 | 3.689665 | 34 | 34 | 22 | |
| Exon_290 | 178599554 | 178599850 | A-band | 297 | 96 | 100 | 100 | 17.0731 | 34.3947 | 21.8 | 2.88716 | 4.72423 | 3.70627 | 34 | 34 | 23 | |
| Exon_291 | 178599146 | 178599445 | A-band | 300 | 93 | 100 | 100 | 17.0731 | 34.3947 | 22.38675 | 2.88716 | 4.72423 | 3.688515 | 36 | 36 | 22 | |
| Exon_292 | 178598748 | 178599062 | A-band | 315 | 94 | 100 | 100 | 17.0731 | 40.1641 | 22.83825 | 2.88716 | 4.72423 | 3.70627 | 40 | 40 | 26 | |
| Exon_293 | 178598506 | 178598654 | A-band | 149 | 95 | 100 | 100 | 20.0197 | 40.1641 | 26.6151 | 2.88716 | 4.83387 | 3.75664 | 35 | 35 | 22 | |
| Exon_294 | 178597908 | 178598058 | A-band | 151 | 96 | 100 | 100 | 20.0197 | 40.1641 | 29.3718 | 2.88716 | 4.83387 | 3.82389 | 33 | 33 | 22 | |
| Exon_295 | 178597538 | 178597819 | A-band | 282 | 100 | 100 | 100 | 20.0197 | 40.1641 | 29.3718 | 2.88716 | 4.83387 | 3.82389 | 33 | 33 | 22 | |
| Exon_296 | 178595507 | 178595809 | A-band | 303 | 92 | 100 | 100 | 20.0197 | 40.1641 | 29.3718 | 2.88716 | 4.83387 | 4.02808 | 39 | 39 | 29 | |
| Exon_297 | 178594344 | 178594646 | A-band | 303 | 90 | 100 | 100 | 20.0197 | 40.1641 | 29.3718 | 2.88716 | 4.83387 | 4.02808 | 39 | 39 | 33 | |
| Exon_298 | 178593961 | 178594242 | A-band | 282 | 90 | 100 | 100 | 20.0197 | 40.1641 | 29.6152 | 3.25779 | 4.83387 | 4.0548 | 35 | 35 | 31 | |
| Exon_299 | 178593568 | 178593867 | A-band | 300 | 89 | 100 | 100 | 21.6197 | 40.1641 | 29.535 | 3.2122 | 4.83387 | 4.05178 | 37 | 37 | 30 | |
| Exon_300 | 178593173 | 178593475 | A-band | 303 | 90 | 100 | 100 | 18.8501 | 40.1641 | 29.6152 | 2.8943 | 4.83387 | 4.05178 | 35 | 35 | 27 | |

Table S2 - TTN exons and metadata

| Exon | hg38 start | hg38 end | band | Coding length | GTEx PSI | DCM PSI | Max PSI | CM: min OR for overlapping windows | CM: max OR for overlapping windows | CM: median OR for overlapping windows | Afib: min OR for overlapping windows | Afib: max OR for overlapping windows | Afib: median OR for overlapping windows | n windows | CM: n sig windows | Afib: n sig windows | Note |
|---|---|---|---|---|---|---|---|---|---|---|---|---|---|---|---|---|---|
| Exon_301 | 178592775 | 178593083 | A-band | 309 | 90 | 100 | 100 | 18.8501 | 40.1641 | 29.535 | 2.8943 | 4.83387 | 4.02808 | 37 | 37 | 28 | |
| Exon_302 | 178592379 | 178592660 | A-band | 282 | 90 | 100 | 100 | 13.4261 | 40.1641 | 26.40855 | 2.8943 | 4.83387 | 4.05179 | 40 | 40 | 31 | |
| Exon_303 | 178591978 | 178592277 | A-band | 300 | 91 | 100 | 100 | 13.4261 | 40.1641 | 25.3603 | 2.8943 | 4.83387 | 4.24776 | 37 | 37 | 31 | |
| Exon_304 | 178591599 | 178591892 | A-band | 294 | 92 | 100 | 100 | 13.4261 | 38.2882 | 22.4227 | 2.8943 | 4.83387 | 4.25786 | 36 | 36 | 30 | |
| Exon_305 | 178588538 | 178591504 | A-band | 2967 | 97 | 100 | 100 | 13.4261 | 35.7961 | 21.289 | 2.8943 | 6.29671 | 4.31336 | 62 | 62 | 53 | |
| Exon_306 | 178587899 | 178588219 | A-band | 321 | 90 | 100 | 100 | 14.369 | 24.9489 | 20.5417 | 2.90686 | 6.29671 | 4.32667 | 37 | 37 | 32 | |
| Exon_307 | 178587516 | 178587800 | A-band | 285 | 91 | 100 | 100 | 14.369 | 24.9489 | 20.5417 | 2.90686 | 6.29671 | 4.32667 | 35 | 35 | 30 | |
| Exon_308 | 178587118 | 178587417 | A-band | 300 | 92 | 100 | 100 | 14.369 | 24.9489 | 20.7037 | 2.90686 | 6.29671 | 4.34598 | 33 | 33 | 28 | |
| Exon_309 | 178586505 | 178586807 | A-band | 303 | 91 | 100 | 100 | 14.3155 | 24.9489 | 20.8566 | 2.90686 | 6.29671 | 4.32667 | 31 | 31 | 26 | |
| Exon_310 | 178585072 | 178585347 | A-band | 276 | 91 | 100 | 100 | 14.3155 | 24.9489 | 21.1313 | 2.90686 | 6.29671 | 4.5002 | 28 | 28 | 23 | |
| Exon_311 | 178584669 | 178584968 | A-band | 300 | 91 | 100 | 100 | 14.3155 | 24.9489 | 21.14135 | 2.90686 | 6.29671 | 4.40639 | 26 | 26 | 21 | |
| Exon_312 | 178584276 | 178584578 | A-band | 303 | 92 | 100 | 100 | 14.3155 | 24.9489 | 21.14135 | 2.90686 | 6.29671 | 4.14565 | 24 | 24 | 19 | |
| Exon_313 | 178583607 | 178583906 | A-band | 300 | 90 | 100 | 100 | 14.3155 | 24.9489 | 21.1313 | 2.90686 | 6.29671 | 4.06377 | 22 | 22 | 17 | |
| Exon_314 | 178582940 | 178583227 | A-band | 288 | 89 | 100 | 100 | 14.3155 | 24.9489 | 20.78015 | 2.90686 | 5.86939 | 3.687335 | 18 | 18 | 13 | |
| Exon_315 | 178582296 | 178582592 | A-band | 297 | 88 | 100 | 100 | 14.3155 | 24.9489 | 20.6227 | 1.59105 | 5.86939 | 3.67839 | 18 | 18 | 12 | |
| Exon_316 | 178581906 | 178582208 | A-band | 303 | 86 | 100 | 100 | 14.3155 | 21.1514 | 20.2229 | 1.58499 | 4.22753 | 3.54933 | 15 | 15 | 8 | |
| Exon_317 | 178581499 | 178581804 | A-band | 306 | 88 | 100 | 100 | 14.3155 | 21.1514 | 20.046 | 1.58499 | 4.06377 | 3.42943 | 13 | 13 | 6 | |
| Exon_318 | 178580322 | 178580609 | A-band | 288 | 91 | 100 | 100 | 14.3155 | 20.2229 | 17.03205 | 1.58499 | 3.69455 | 3.33977 | 8 | 8 | 1 | |
| Exon_319 | 178579939 | 178580229 | A-band | 291 | 92 | 100 | 100 | 14.3155 | 18.4953 | 15.386 | 1.58499 | 4.28671 | 3.168145 | 6 | 6 | 2 | |
| Exon_320 | 178579561 | 178579848 | A-band | 288 | 92 | 100 | 100 | 11.2597 | 23.0603 | 15.1058 | 1.58499 | 6.0023 | 4.24489 | 35 | 33 | 28 | |
| Exon_321 | 178578806 | 178579393 | A-band | 588 | 92 | 100 | 100 | 10.8393 | 23.0603 | 14.6927 | 2.87992 | 6.0023 | 4.14245 | 35 | 31 | 27 | |
| Exon_322 | 178578611 | 178578715 | A-band | 105 | 91 | 100 | 100 | 10.8393 | 23.0603 | 14.9366 | 2.87992 | 6.0023 | 4.26347 | 31 | 27 | 26 | |
| Exon_323 | 178577988 | 178578185 | A-band | 198 | 91 | 100 | 100 | 10.8393 | 23.0603 | 14.9366 | 2.87992 | 6.0023 | 4.26347 | 31 | 27 | 25 | |
| Exon_324 | 178577602 | 178577898 | A-band | 297 | 91 | 100 | 100 | 10.8393 | 23.0603 | 14.6428 | 2.86634 | 6.0023 | 4.19367 | 34 | 30 | 23 | |
| Exon_325 | 178576923 | 178577510 | A-band | 588 | 93 | 100 | 100 | 10.8393 | 23.0603 | 14.657 | 2.65242 | 5.88782 | 3.656875 | 38 | 35 | 20 | |
| Exon_326 | 178576529 | 178576831 | A-band | 303 | 94 | 100 | 100 | 10.8393 | 23.0603 | 14.3486 | 2.65242 | 5.58785 | 3.253925 | 44 | 41 | 17 | |
| Exon_327 | 178559311 | 178576416 | A-band | 17106 | 95 | 100 | 100 | 10.8393 | 36.4516 | 20.495 | 1.89523 | 7.9228 | 3.27959 | 175 | 170 | 74 | |
| Exon_328 | 178558341 | 178558637 | A-band | 297 | 96 | 100 | 100 | 16.6534 | 31.287 | 23.865 | 2.29228 | 3.34496 | 2.82042 | 41 | 41 | 0 | |
| Exon_329 | 178557648 | 178558235 | A-band | 588 | 95 | 100 | 100 | 16.6534 | 31.287 | 23.441 | 2.11971 | 3.34496 | 2.74991 | 43 | 43 | 0 | |
| Exon_330 | 178557253 | 178557555 | A-band | 303 | 96 | 100 | 100 | 16.6534 | 31.287 | 23.346 | 2.11971 | 3.34496 | 2.82592 | 42 | 42 | 0 | |
| Exon_331 | 178556848 | 178557144 | A-band | 297 | 96 | 100 | 100 | 16.6534 | 31.287 | 23.4209 | 2.11971 | 3.34496 | 2.82042 | 43 | 43 | 0 | |
| Exon_332 | 178554865 | 178555152 | A-band | 288 | 96 | 100 | 100 | 16.6534 | 31.287 | 23.6526 | 2.11971 | 3.34496 | 2.83266 | 39 | 39 | 0 | |
| Exon_333 | 178554453 | 178554752 | A-band | 300 | 96 | 100 | 100 | 16.6534 | 31.287 | 23.6526 | 2.11971 | 3.34496 | 2.83266 | 41 | 41 | 0 | |
| Exon_334 | 178553914 | 178554216 | A-band | 303 | 97 | 100 | 100 | 19.9165 | 31.287 | 24.9079 | 2.11971 | 3.93393 | 2.85598 | 39 | 39 | 1 | |
| Exon_335 | 178553502 | 178553807 | A-band | 306 | 97 | 100 | 100 | 19.9165 | 31.287 | 24.9079 | 2.11971 | 3.93393 | 2.866285 | 38 | 38 | 1 | |
| Exon_336 | 178551630 | 178553396 | A-band | 1767 | 97 | 100 | 100 | 8.16756 | 31.287 | 23.75105 | 2.11971 | 3.93393 | 2.84432 | 46 | 39 | 2 | |
| Exon_337 | 178550967 | 178551260 | A-band | 294 | 98 | 100 | 100 | 8.16756 | 25.3843 | 18.18615 | 2.11971 | 3.93393 | 2.910105 | 32 | 24 | 2 | |
| Exon_338 | 178549986 | 178550273 | A-band | 288 | 98 | 100 | 100 | 0.365089 | 25.3843 | 13.81945 | 2.45265 | 3.93393 | 3.00211 | 32 | 21 | 2 | |
| Exon_339 | 178549570 | 178549869 | A-band | 300 | 98 | 100 | 100 | 0.364915 | 25.2517 | 11.3161 | 2.14712 | 3.93393 | 2.95589 | 34 | 15 | 2 | |
| Exon_340 | 178547407 | 178549473 | A-band | 2067 | 98 | 100 | 100 | 0.364915 | 23.3128 | 6.55606 | 2.14712 | 3.93393 | 2.85506 | 48 | 12 | 3 | |
| Exon_341 | 178547003 | 178547305 | A-band | 303 | 98 | 100 | 100 | 0.364915 | 11.2697 | 4.747475 | 2.14712 | 4.22213 | 2.804505 | 26 | 0 | 2 | |
| Exon_342 | 178546600 | 178546905 | A-band | 306 | 98 | 100 | 100 | 0.364915 | 11.2697 | 5.640085 | 2.14712 | 4.22213 | 2.817545 | 26 | 0 | 4 | |
| Exon_343 | 178546212 | 178546502 | A-band | 291 | 98 | 100 | 100 | 0.364915 | 11.6029 | 5.843785 | 2.60621 | 4.22213 | 2.85506 | 26 | 0 | 4 | |
| Exon_344 | 178545820 | 178546116 | A-band | 297 | 99 | 100 | 100 | 0.364919 | 18.3573 | 7.27861 | 2.60621 | 4.22213 | 3.071955 | 30 | 6 | 10 | |
| Exon_345 | 178545388 | 178545693 | A-band | 306 | 99 | 100 | 100 | 4.13793 | 22.9252 | 8.58632 | 2.60621 | 4.3107 | 3.45357 | 31 | 9 | 13 | |
| Exon_346 | 178544201 | 178544506 | A-band | 306 | 99 | 100 | 100 | 4.13793 | 22.9252 | 11.41105 | 2.60621 | 5.18207 | 3.73532 | 34 | 15 | 19 | |
| Exon_347 | 178543834 | 178544115 | A-band | 282 | 99 | 100 | 100 | 5.81665 | 24.8057 | 18.3573 | 2.82221 | 5.88747 | 4.08128 | 35 | 23 | 27 | |
| Exon_348 | 178543069 | 178543662 | A-band | 594 | 99 | 100 | 100 | 7.97999 | 24.8057 | 19.8586 | 3.1769 | 6.71368 | 4.61238 | 41 | 33 | 37 | |
| Exon_349 | 178542662 | 178542949 | A-band | 288 | 99 | 100 | 100 | 11.0195 | 26.9509 | 20.5275 | 3.1769 | 8.37931 | 4.70381 | 37 | 34 | 34 | |
| Exon_350 | 178542264 | 178542563 | A-band | 300 | 100 | 100 | 100 | 16.4263 | 26.9509 | 20.5989 | 3.73264 | 8.37931 | 5.05803 | 37 | 37 | 37 | |
| Exon_351 | 178541282 | 178541584 | A-band | 303 | 98 | 100 | 100 | 16.4263 | 26.9509 | 20.5989 | 3.73264 | 8.37931 | 5.48191 | 37 | 37 | 37 | |
| Exon_352 | 178540068 | 178540370 | A-band | 303 | 100 | 100 | 100 | 14.1546 | 26.9509 | 20.5989 | 4.3107 | 8.37931 | 5.88747 | 35 | 35 | 35 | |
| Exon_353 | 178539382 | 178539966 | A-band | 585 | 100 | 100 | 100 | 11.105 | 26.9509 | 18.2993 | 4.44725 | 8.37931 | 5.68943 | 46 | 43 | 46 | |
| Exon_354 | 178538946 | 178539251 | A-band | 306 | 100 | 100 | 100 | 11.105 | 26.9509 | 18.11735 | 4.44725 | 8.37931 | 5.85553 | 40 | 37 | 40 | |
| Exon_355 | 178538540 | 178538839 | A-band | 300 | 100 | 100 | 100 | 11.105 | 26.9509 | 18.23285 | 3.75102 | 8.37931 | 5.703755 | 44 | 41 | 44 | |
| Exon_356 | 178537342 | 178537917 | A-band | 576 | 100 | 100 | 100 | 11.105 | 26.9509 | 18.1672 | 1.89946 | 8.37931 | 5.68943 | 45 | 42 | 41 | |
| Exon_357 | 178536938 | 178537243 | A-band | 306 | 99 | 100 | 100 | 11.105 | 23.9627 | 17.5378 | 1.89946 | 6.83382 | 4.755845 | 36 | 33 | 29 | |
| Exon_358 | 178535982 | 178536575 | A-band | 594 | 99 | 100 | 100 | 11.105 | 26.7342 | 18.1672 | 1.89946 | 6.83382 | 4.44725 | 41 | 38 | 26 | |
| Exon_359 | 178530241 | 178535849 | M-band | 5609 | 100 | 100 | 100 | 0.364296 | 26.7342 | 11.5103 | 0.578858 | 4.87568 | 2.84806 | 89 | 44 | 27 | There are no associated windows past position chr2:178531887, corresponding to amino acid position 34909 |
| Exon_360 | 178529960 | 178530116 | M-band | 157 | 99 | 100 | 100 | 0.364364 | 0.364476 | 0.36441 | 0.611103 | 1.26366 | 0.813742 | 19 | 0 | 0 | |
| Exon_361 | 178528528 | 178529219 | M-band | 692 | 99 | 100 | 100 | 0.364364 | 0.364476 | 0.3644135 | 0.611103 | 1.26366 | 0.836232 | 16 | 0 | 0 | |
| Exon_362 | 178528274 | 178528427 | M-band | 154 | 100 | 100 | 100 | 0.364202 | 0.364593 | 0.364446 | 0.847808 | 1.19209 | 0.963987 | 19 | 0 | 0 | |
| Exon_363 | 178527446 | 178527748 | M-band | 303 | 99 | 99 | 99 | 0.364202 | 0.364593 | 0.364458 | 0.915926 | 1.08871 | 0.97263 | 14 | 0 | 0 | |
| Exon_364 | 178525989 | 178527307 | M-band | 296 | 100 | 100 | 100 | 0.364368 | 0.364593 | 0.364545 | 0.931687 | 1.08871 | 1.08031 | 8 | 0 | 0 | |

*Exon_48 is not part of the canonical transcript and was not included in this analysis

**Table S3. UKB and HNP cohort demographics and phecode summary**

| EHR depth | Cohort | n | % Female | Mean (std) age in 2021 | % CM (Phecode 425.1) | % Afib (Phecode 427.2) | Mean (std) phecode range in years |
|---|---|---|---|---|---|---|---|
| Any ICD hx | UKB450K | 461,738 | 54.5%(251,517) | 70(8) | 0.7%(3,297) | 7.5% (34,407) | 25 (18) |
| | HNP25K | 32,030 | 67.4% (21,589) | 53(17) | 1.4% (441) | 4.6% (1,489) | 7 (4) |
| >1 year ICD hx | UKB450K | 388,070 | 54.8%(213,011) | 70(8) | 0.8%(3,013) | 8.1%(31,517) | 27 (17) |
| | HNP25K | 25,493 | 68.9% (17,556) | 53(17) | 1.4%(362) | 4.9%(1,261) | 8 (4) |

CM and Afib rates are in line with other estimates from cohort-based studies (3,26). For % Female, % CM, and % Afib columns raw counts are noted after percentages. Mean phecode range in years is meant to provide a quantative representation of the average depth of EHR data per participant

**Table S4. Breakdown of temprotal diagnosis patterning of individuals with both atrial fibrillation (Afib) and cardiomyopathy (CM) phecodes in the UKB450k**

| Cases by *TTN*tv genotype status | Afib before CM | Afib and CM concurrent | CM before Afib |
|---|---|---|---|
| hiPSI carriers (n=89) | 42% | 30% | 28% |
| non-carriers (n=1373) | 45% | 30% | 25% |
| loPSI carriers (n=12) | 25% | 16% | 59% |

Number of individuals in each genotypic group is indicated next to the carrier designation. CM and Afib phecodes are compared at year level resolution. Percentages are rounded to the nearest whole number. Afib is most often diagnosed before CM.
